# Theory-based self-management interventions for stroke survivors: a systematic review and meta-analysis

**DOI:** 10.64898/2026.03.02.26346812

**Authors:** Guangyan Meng, Yuping Chen, Mengting Dai, Qirong Chen, Siyuan Tang

## Abstract

**Background:** Self-management is essential for stroke survivors to maintain a healthy lifestyle and reduce recurrence risk. Although theory-based self-management interventions are widely recommended, the theoretical frameworks underpinning them and their comparative effectiveness remain unclear.

**Aims:** To systematically identify the theories, models, and frameworks (TMFs) used in self-management interventions for stroke survivors, to explore how they guide interventions, and evaluate their effectiveness on self-management behaviors and self-efficacy.

**Methods:** PubMed, Embase, Web of Science, ProQuest Health & Medical Collection and the Cochrane Library were searched from inception to July 15, 2025. Randomized controlled trials or quasi-experimental studies evaluating theory-based self-management interventions for stroke survivors were included. Two reviewers independently screened studies, extracted data, and assessed risk of bias (Cochrane RoB 2.0). Meta-analyses were performed using random-effects models.

**Results:** From 11,495 records, 32 studies with 3,212 participants were included. Sixteen distinct TMFs were identified; self-efficacy theory was most frequent (13/32), followed by social cognitive theory (6/32). All TMFs were middle-range theories. Meta-analysis showed TMFs-based interventions significantly improved self-management behaviors (SMD = 4.26, 95%CI: 0.20–8.31, I² = 98.2%) and self-efficacy (SMD = 0.60, 95%CI: 0.32–0.88, I² = 72.8%). However, the effect for behaviors is likely inflated due to extreme heterogeneity and theoretical diversity. Theory-specific analysis of self-efficacy theory (k = 8) confirmed significant effects on self-efficacy (SMD = 0.64, 95%CI: 0.21–1.08).

**Conclusions:** This review identified 16 distinct theoretical models; self-efficacy theory was most frequently applied, followed by social cognitive theory.

Theory-based interventions significantly improved self-management behaviours and self-efficacy.

## 1. Introduction

Stroke is a major public health burden and the second leading cause of death worldwide, with approximately 5.5 million annual fatalities.^[1–3]^ As populations age, incidence is expected to rise, making secondary prevention critical.^[4]^ Nearly 90% of the global stroke burden is linked to modifiable lifestyle risk factors,^[5]^ underscoring the significance for effective secondary prevention. Enhancing self-management among stroke survivors represents a promising approach to sustaining long-term risk factor control and improving health outcomes.^[6, 7]^

Self-management was defined by the National Clinical Guideline for Stroke in the UK as the *“actions and confidence of individuals to manage the medical and emotional aspects of their condition in order to maintain or create new life roles”*.^[8]^ In other words, this definition conceptualizes self-management as comprising both concrete behaviors and the confidence to perform them. For stroke survivors, self-management behavior is defined as the individual’s capacity to exert control over their condition through actions such as medication management, self-care, exercise, and lifestyle adjustments.^[9]^ The adoption of such behaviors is crucial for mitigating future stroke risk.^[10]^ In healthcare, this confidence, termed self-efficacy, is more precisely defined as the belief in one’s ability to carry out the specific behaviors necessary to reach a desired goal,^[11–13]^ and to overcome perceived and actual barriers.^[14]^ Consequently, this review evaluates the effectiveness of interventions by examining both self-management behaviors and the self-efficacy that underlies them as primary outcomes.

Appropriate theoretical foundations are crucial for developing effective, replicable, and explainable interventions.^[15]^ However, despite this consensus, the application of theory in stroke self-management research remains inconsistent. A prior review by Lynch et al., noted that nearly half (45%, n=20) of included interventions lacked any theoretical basis.^[16]^ Even when theories are used, they are applied heterogeneously: across diverse theoretical traditions (e.g., cognitive, motivational, self-regulatory) and with varying degrees of fidelity in operationalization. This heterogeneity makes it difficult to discern which theoretical approaches are most effective for promoting self-management in stroke survivors.

Preliminary evidence suggests the potential of theory-guided approaches. For instance, interventions have drawn from the Protection Motivation Theory (focusing on threat and coping appraisals) to improve self-management ability,^[17]^ while others have integrated the Health Belief Model and Theory of Planned Behavior targeting perceptions and behavioral intentions) to influence lifestyle changes.^[18]^ Additionally, frameworks like Goal Attainment Theory^[19]^ and the Theory of Planned Behavior alone^[20]^ have been utilized to enhance self-management and adherence. While promising, these examples illustrate a fragmented landscape: studies employ distinct theories, often without comparison or a clear rationale for selecting one framework over another. Consequently, it remains unclear whether certain theoretical models (or components thereof) are better suited to address the specific challenges of stroke self-management.

To advance the field, a systematic synthesis is needed to map the theoretical landscape and evaluate comparative effectiveness. Therefore, we seek to answer the following questions: (1) What are the characteristics of the included studies? (2) What TMFs have been used to support self-management interventions for stroke survivors, what are their characteristics, and how were they operationalized? (3) Which TMFs are more effective in improving self-management behaviors and self-efficacy (primary outcomes) and other key health outcomes among stroke survivors?

## 2. Methods

### 2.1 Protocol and Guidance

This systematic review and meta-analysis was performed according to Preferred Reporting Items for Systematic Reviews and Meta-Analyses (PRISMA) guidelines.^[21]^ Our protocol is registered on PROSPERO (CRD420251075002).

### 2.2 Search Strategy

We conducted a literature search in the PubMed, Embase, Web of Science, ProQuest Health & Medical Collection and The Cochrane Library databases from database inception to July 15, 2025. Stroke, self-management, intervention and theory, model or framework were key terms in the literature research. Our search strategies included these terms’ Medical Subject Headings (Me SH) and relevant synonyms to expand the title, abstract, and keyword search in the databases. Full strategies for the databases are available in e Table 1 in supplement.

**Table 1.**
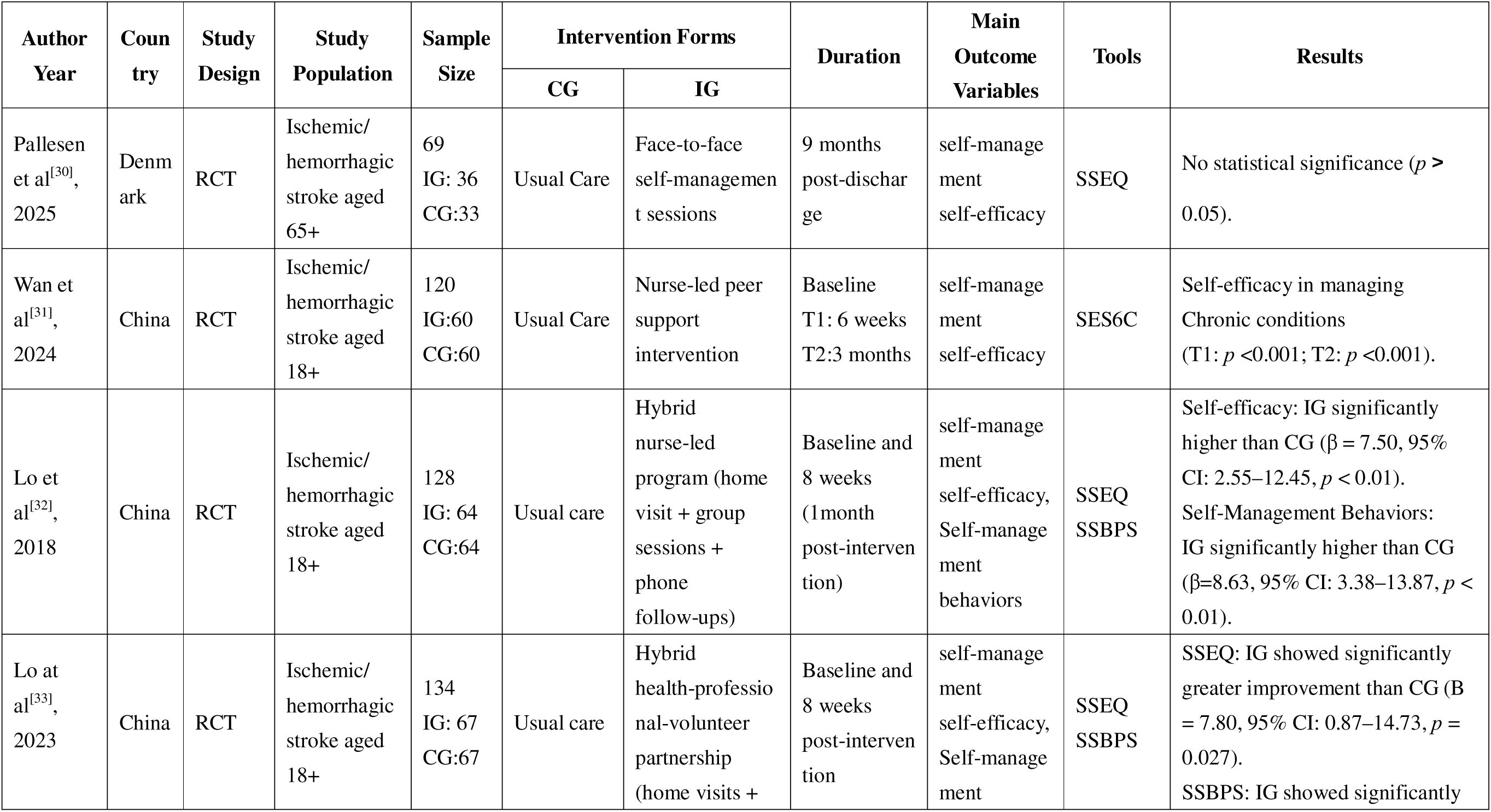
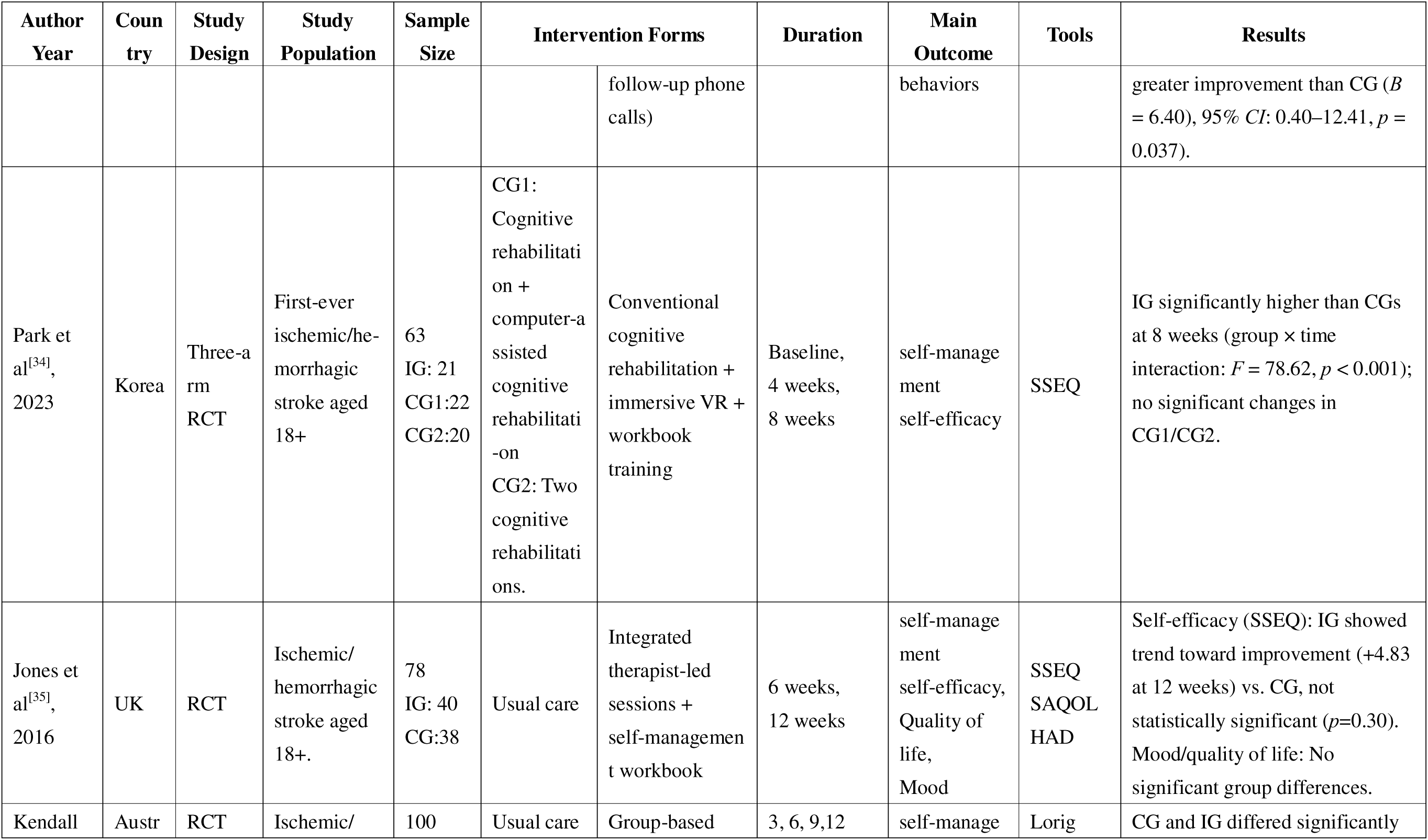
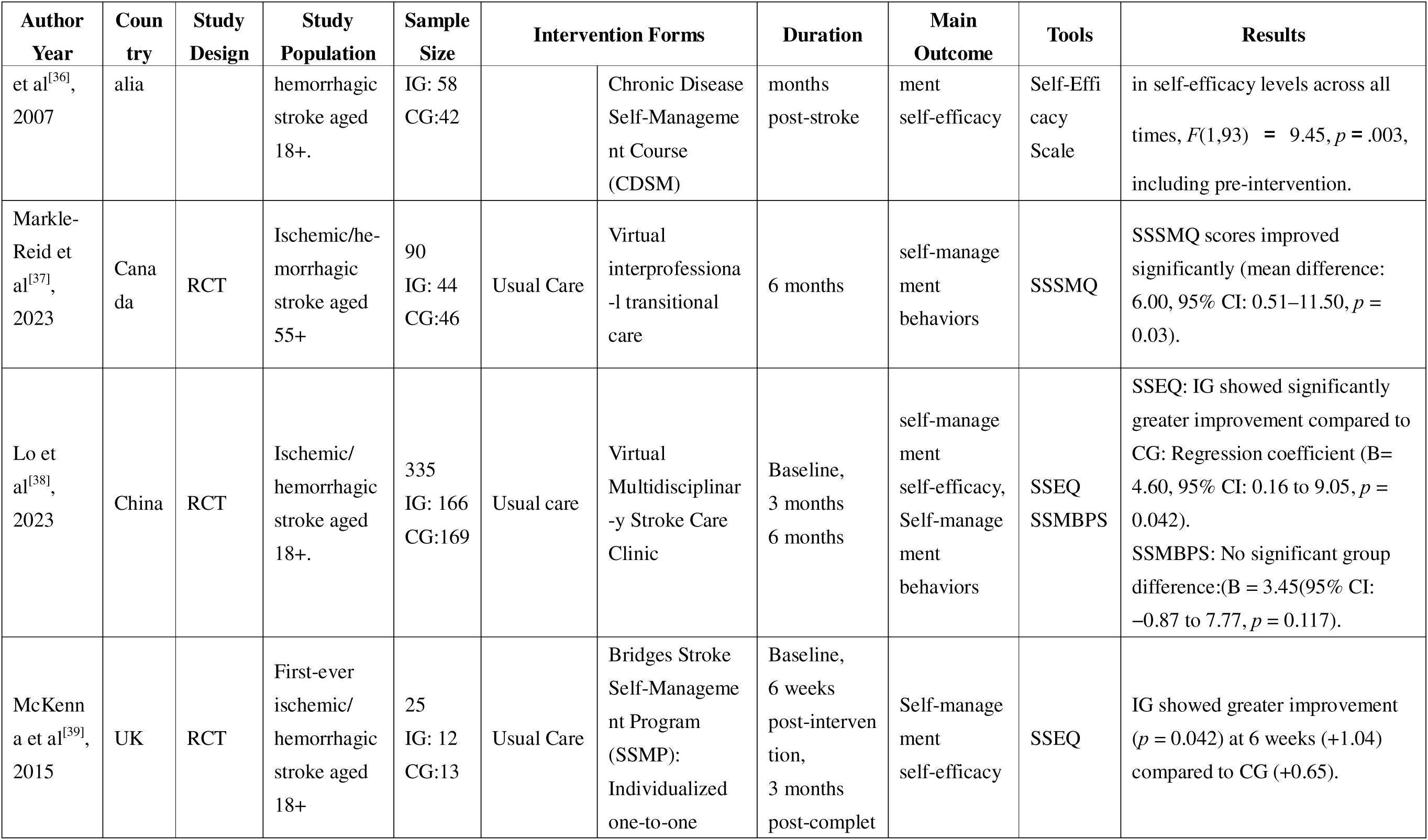
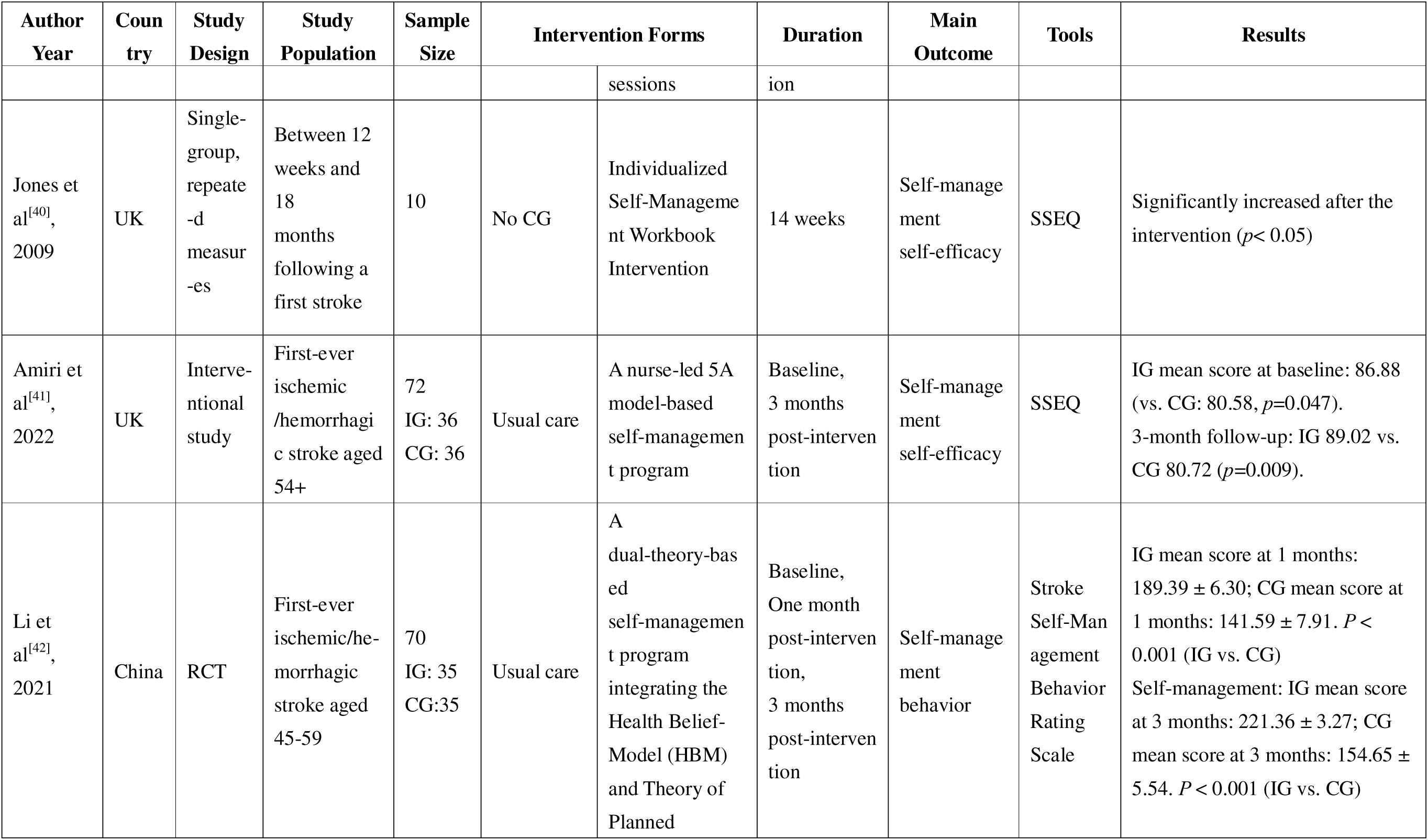
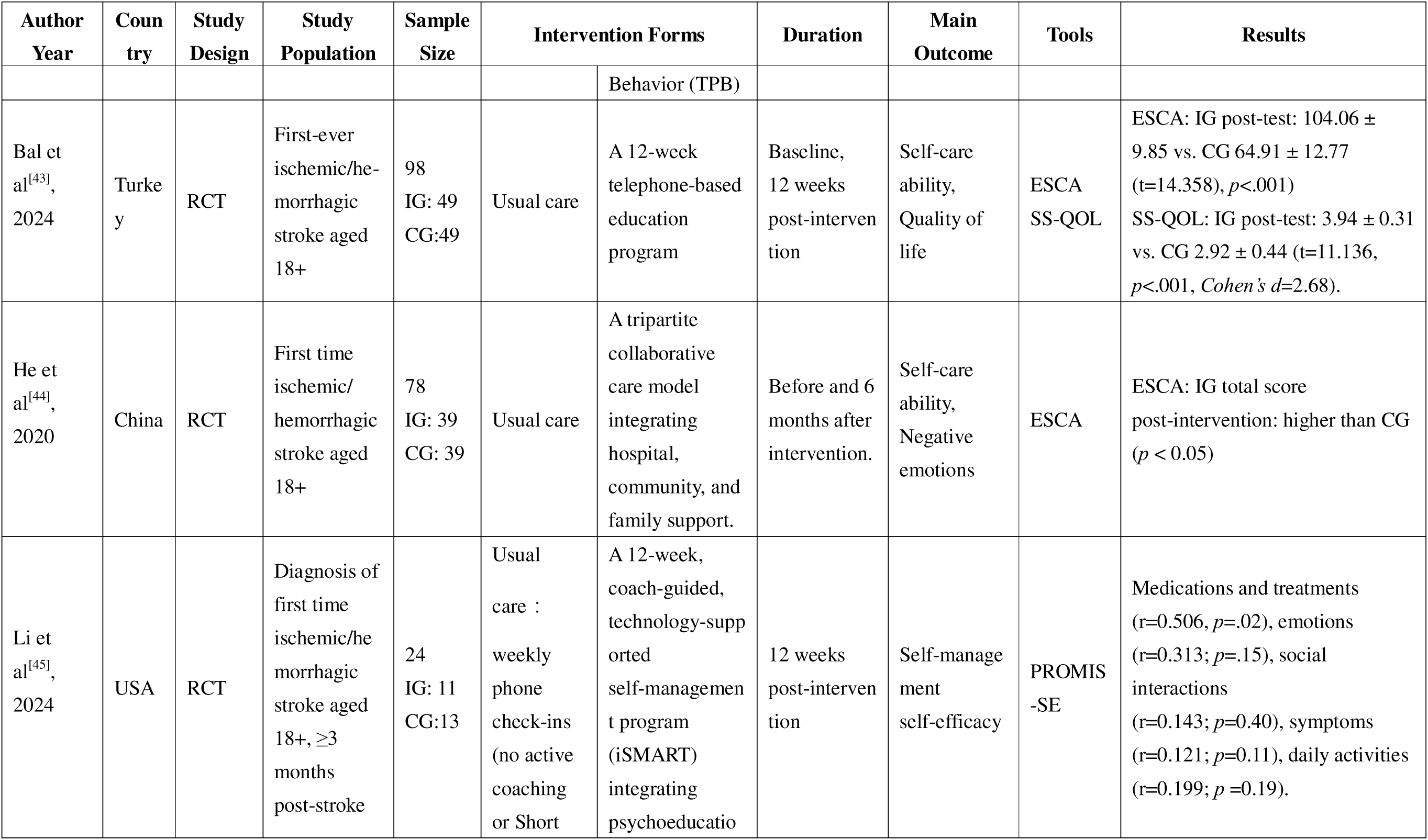
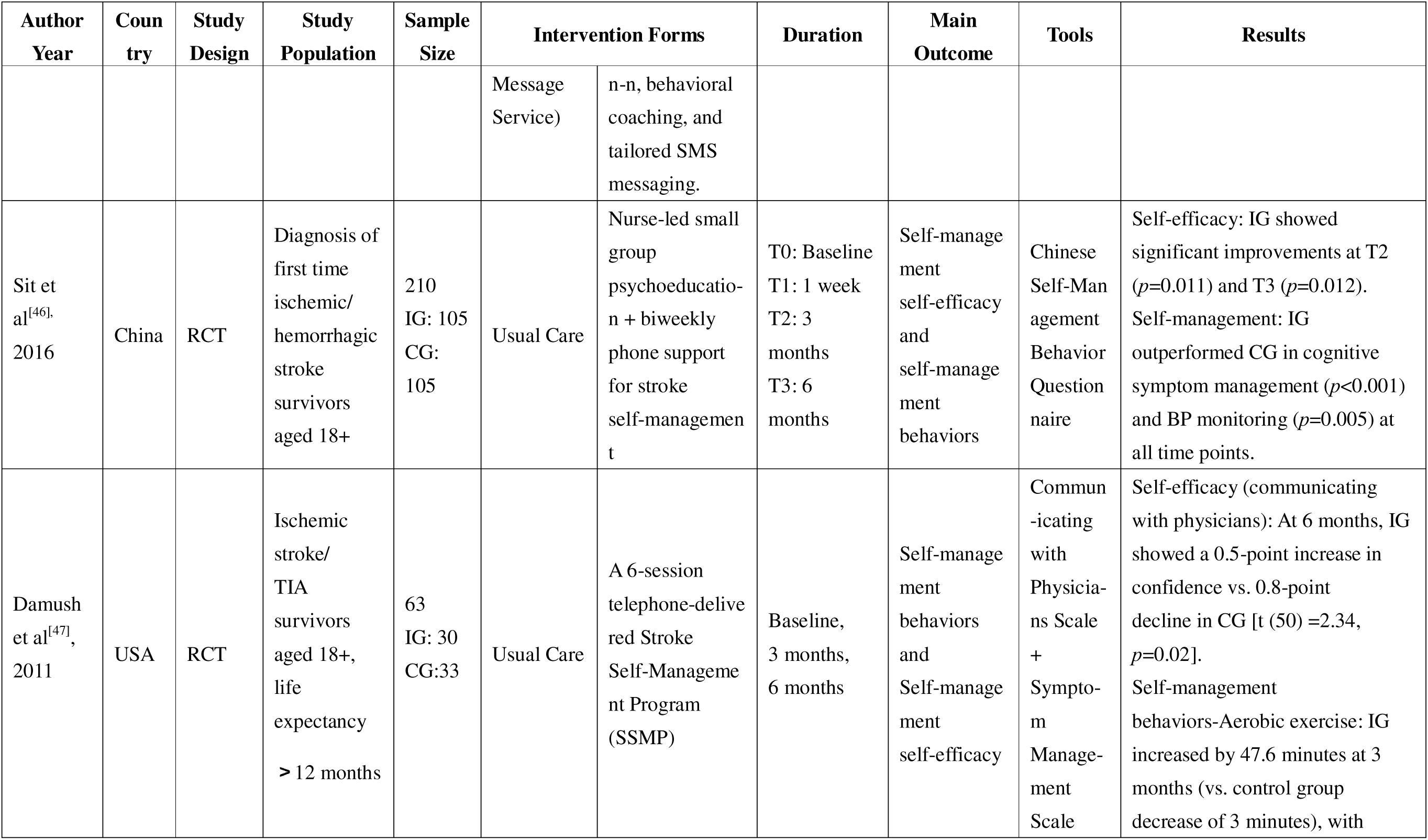
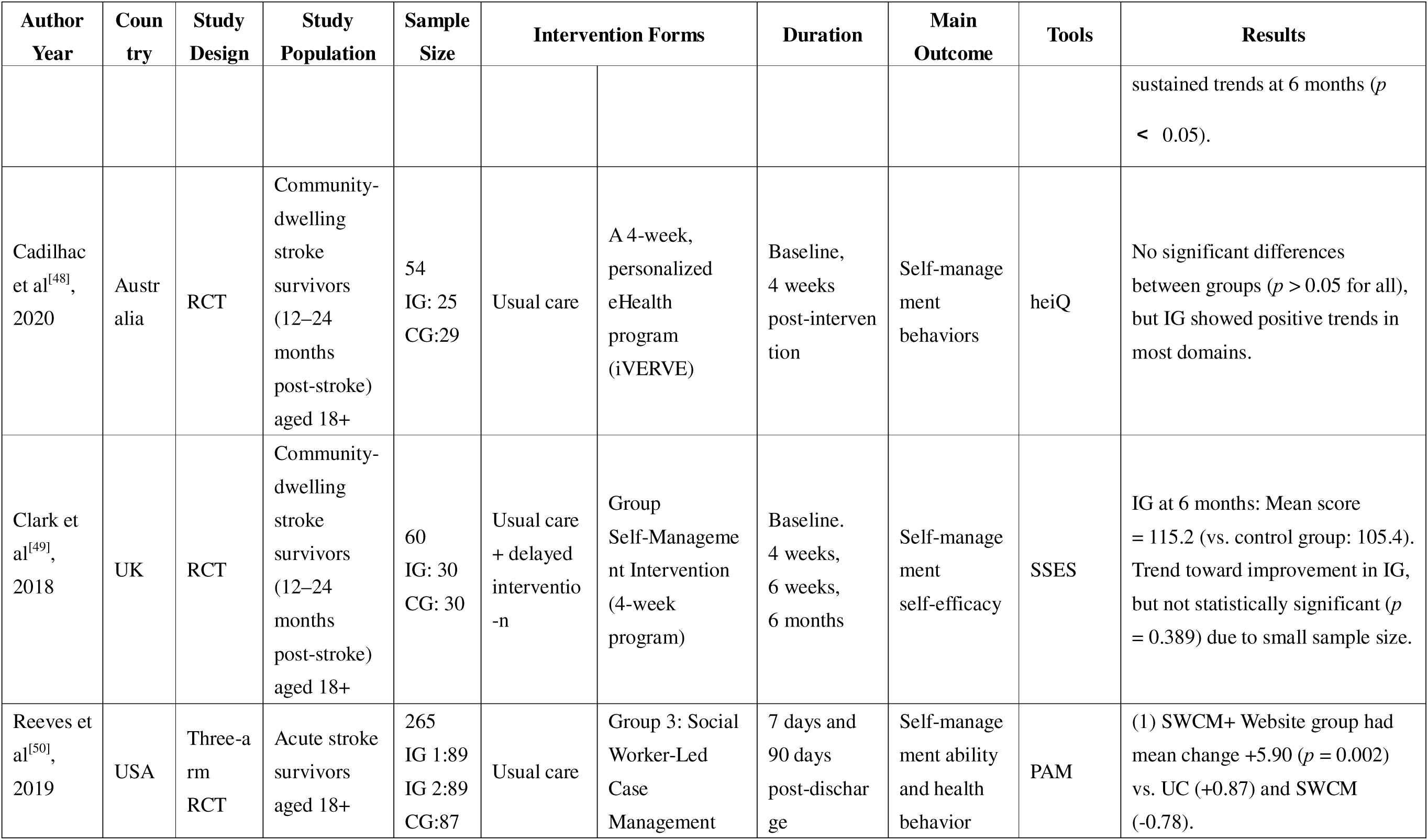
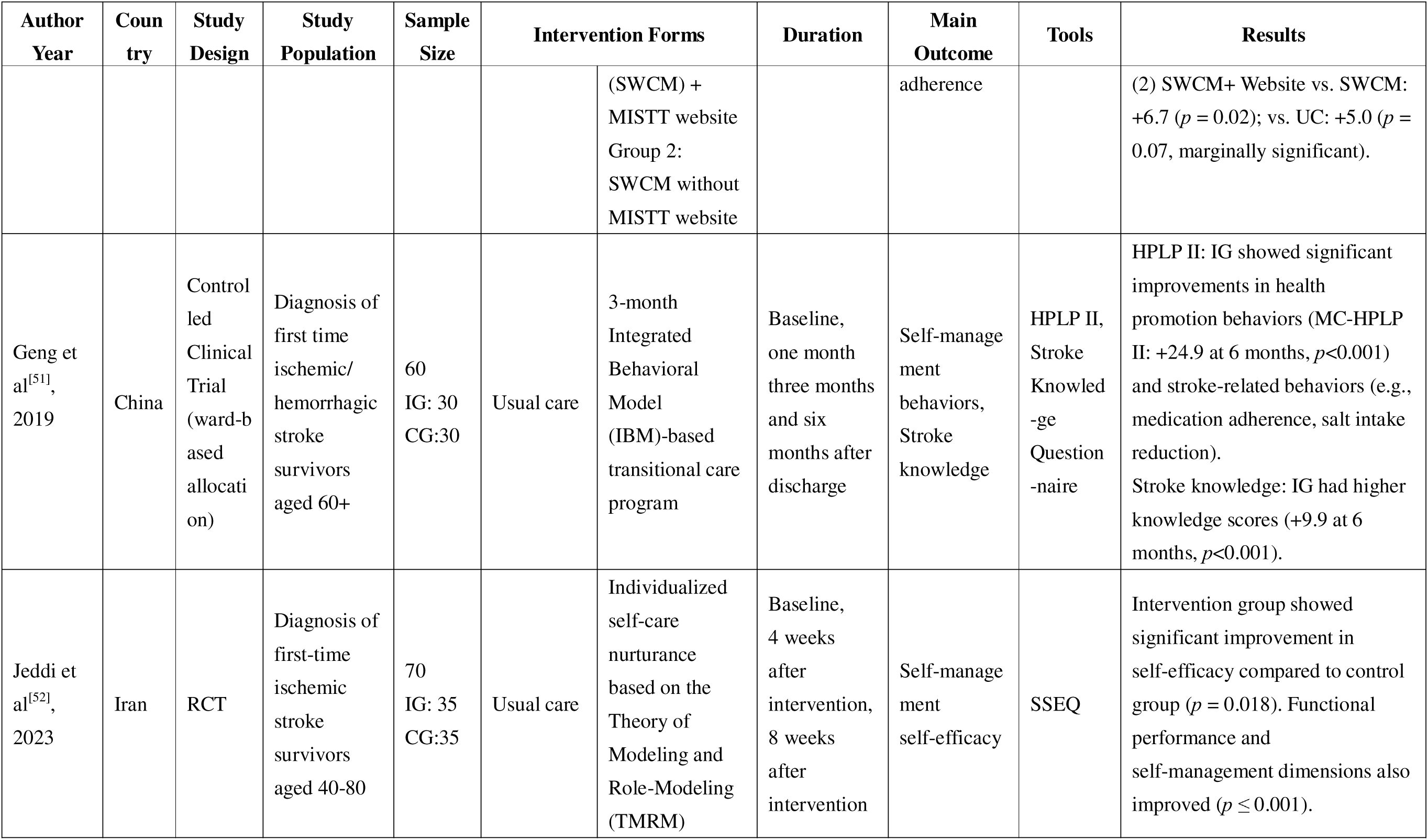
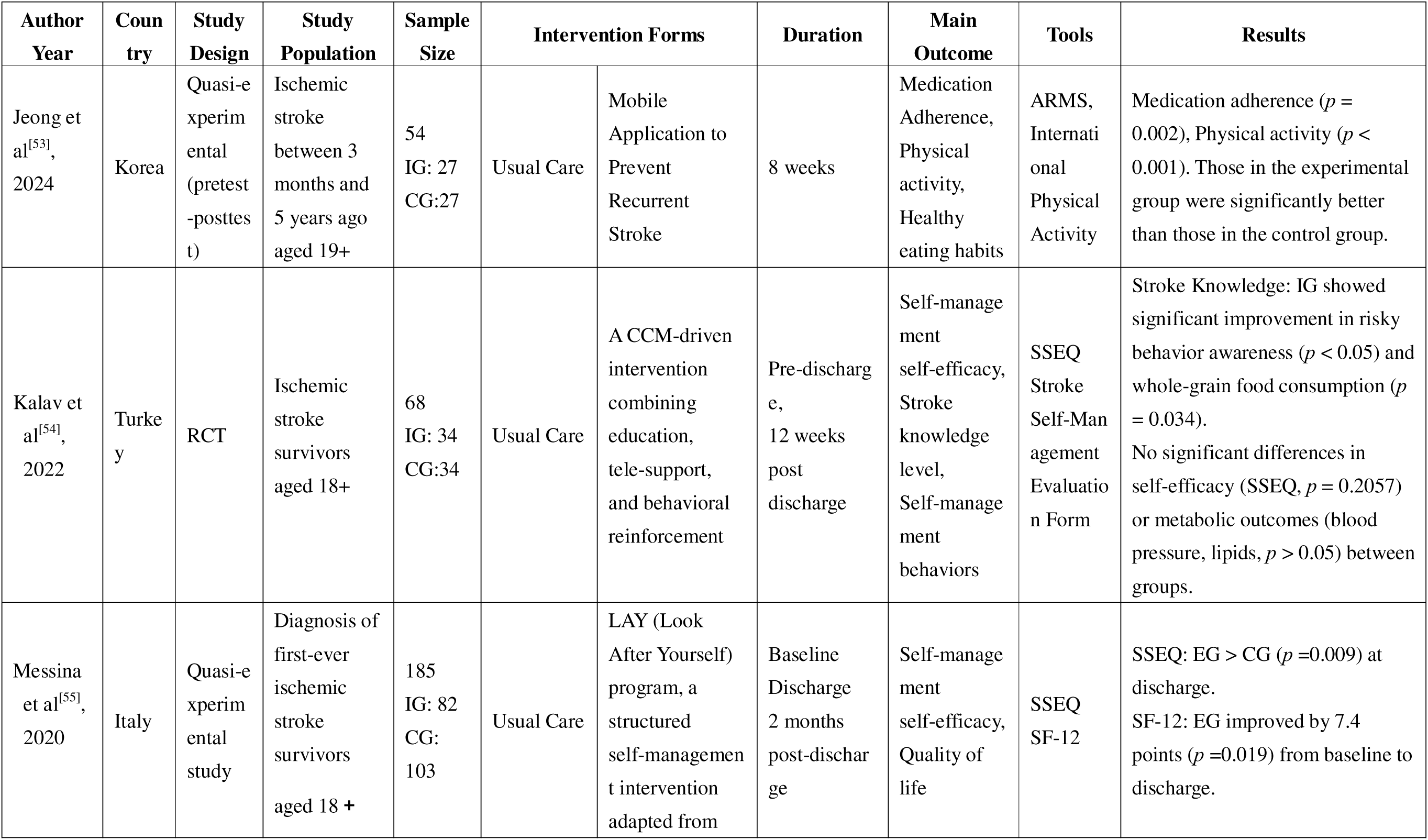
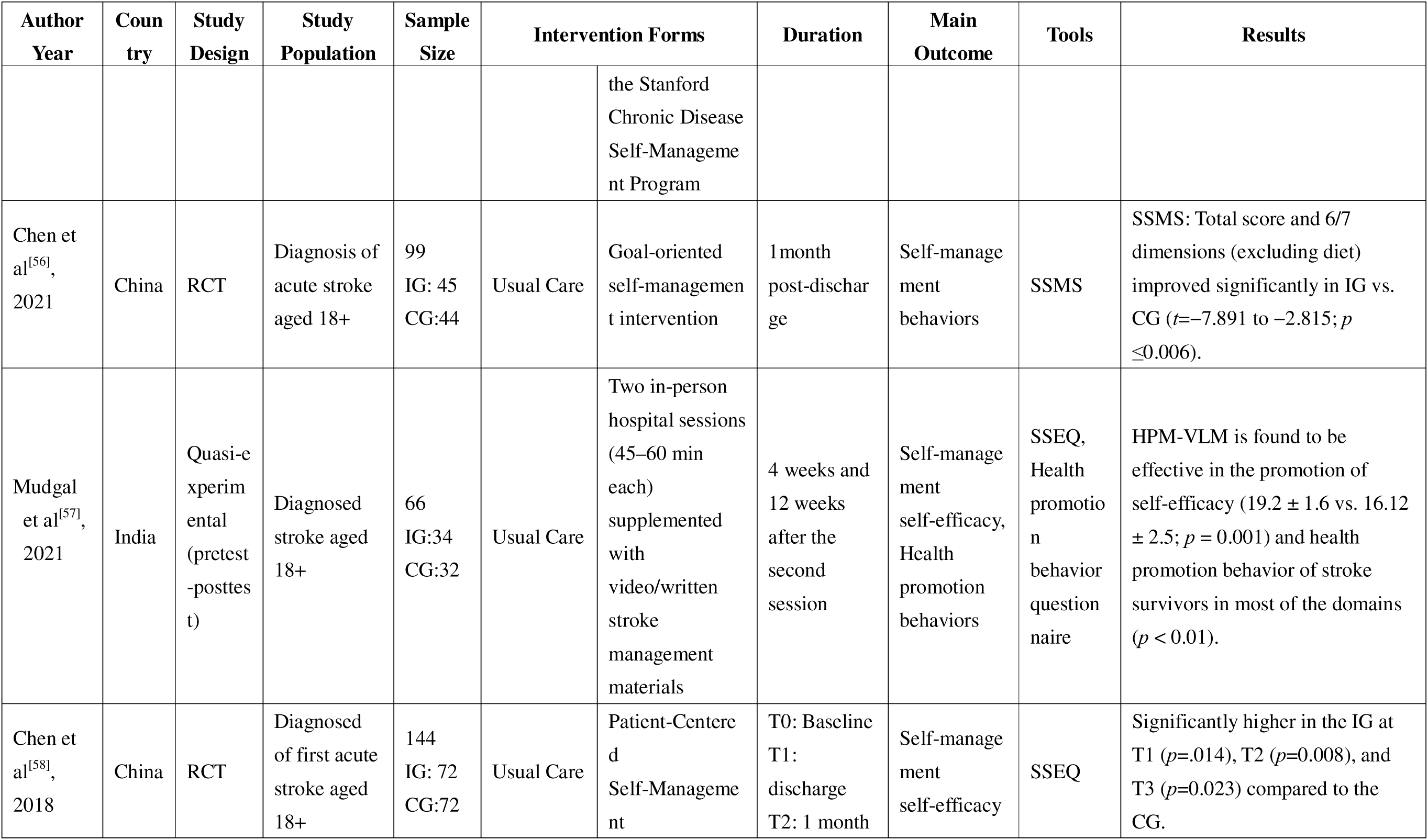
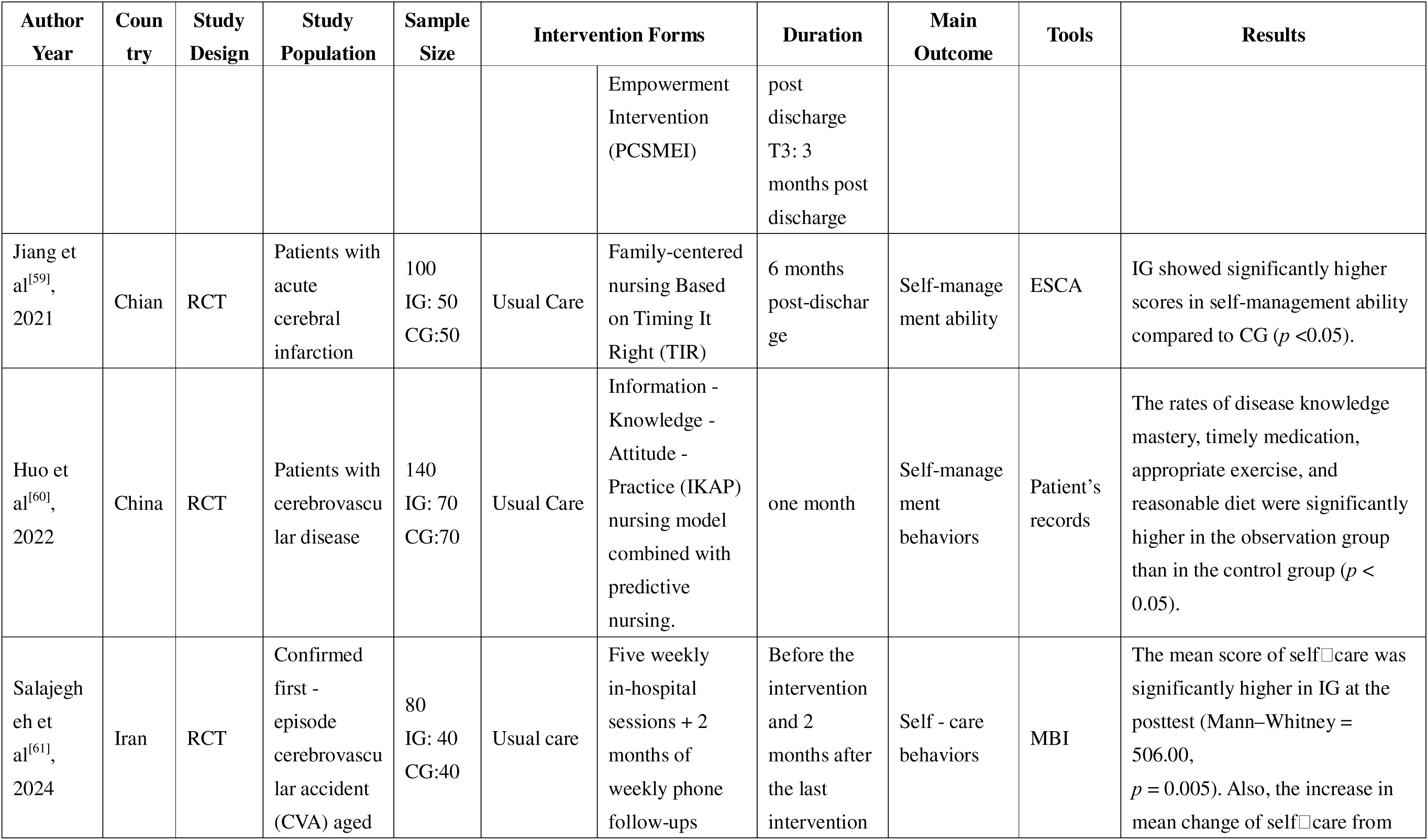
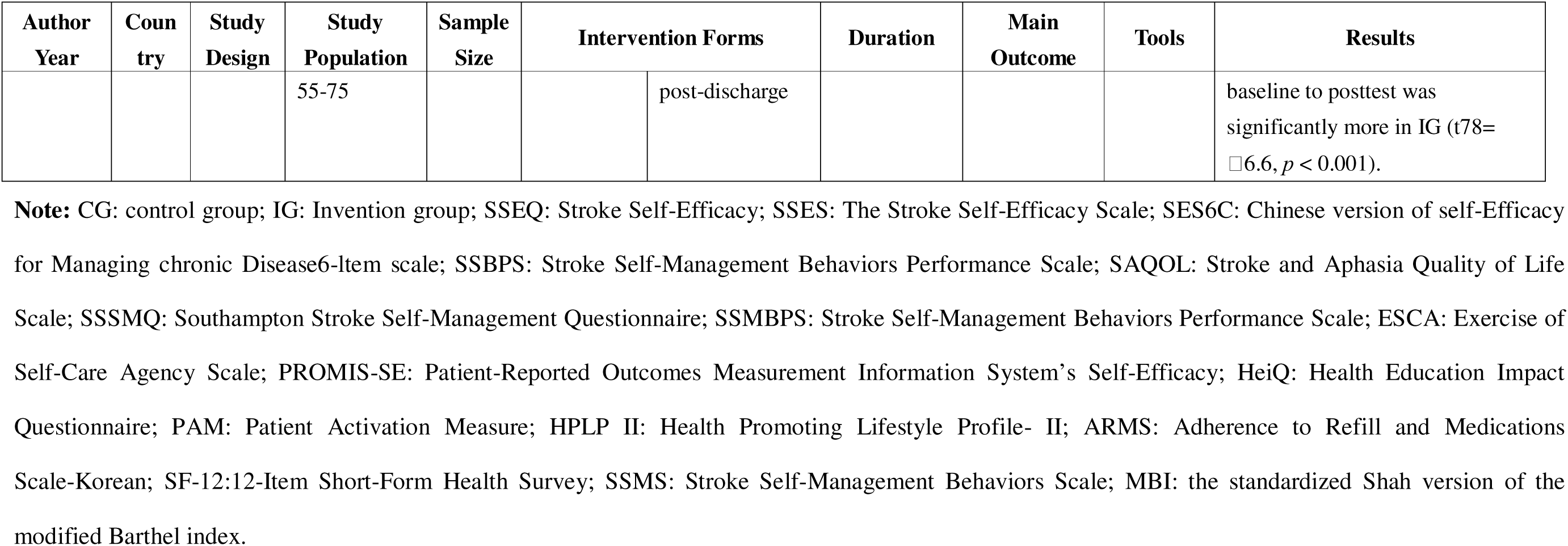
Information of the included studies.

### 2.3 Study Selection

Inclusion criteria: (1) Study population were stroke survivors; (2) Study employs randomized controlled trials (RCTs) or quasi-experimental designs (e.g., non-randomized controlled trials); (3) Interventions explicitly described as being based on or guided by at least one formal theory, model, or framework (TMF). The TMF must be named and its role in intervention design described; (4) Studies reporting data on at least one of our predefined primary outcomes (self-management behaviors or self-efficacy) or key secondary outcomes (e.g., health-related quality of life); (5) Published in English, with no restriction on publication year.

Exclusion criteria:(1) Full text unavailable; (2) Not stroke survivors; (3) Not interventional studies (e.g., reviews, editorials, protocols, qualitative-only studies); (4) Studies not reporting any relevant outcome data. This selection process is shown in Figure 1.

**Figure 1.**
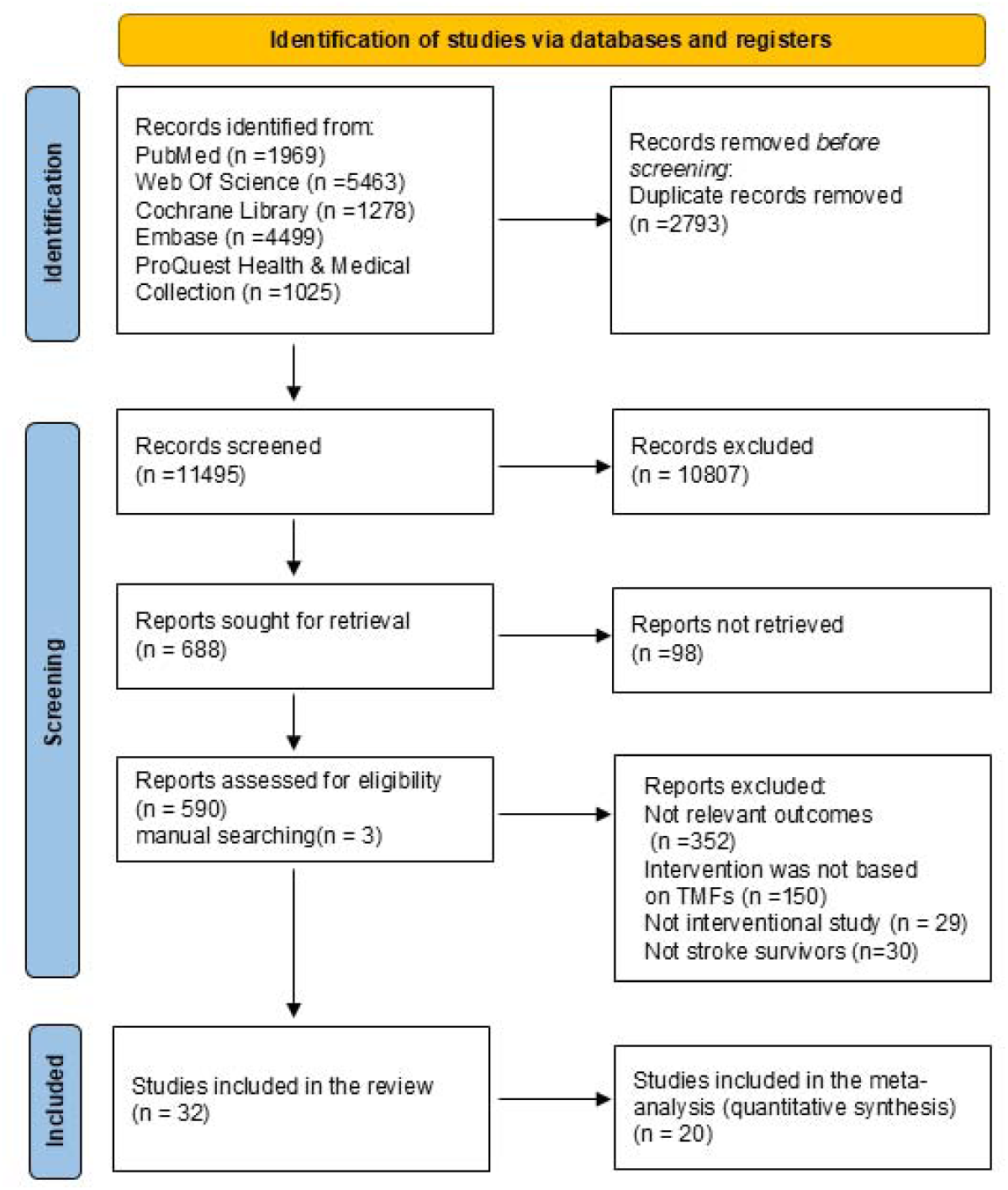
Flow diagram of literature search and study selection process.

Titles and abstracts were exported to Covidence, where they were screened independently by two reviewers. Full-text articles were reviewed independently by two reviewers. Disagreements were resolved through discussions. If no agreement can be reached, consult a third researcher to resolve the differences.

### 2.4 Data Extraction and Synthesis

To address the three research questions, two reviewers independently extracted data. Any discrepancies in screening or extraction were resolved through discussion between the two reviewers. If consensus could not be reached, a third reviewer was consulted to make a final decision. To address research question one (characteristics of the included studies), we constructed a structured extraction Table 1, which captured the following key study characteristics: author(s) and publication year, country of origin, study design, participant population details, sample size for intervention and control groups, form and duration of the intervention, primary and secondary outcome variables, measurement tools employed, and results. To answer question two (which theories, models, or frameworks informed the interventions and how they were applied), we created a structured table (e Table 2). To capture how the TMFs were applied, we developed a extraction table (e Table 3). TMF scope and type were classified using Fawcett and DeSanto-Madeya’s framework for nursing theory analysis and evaluation, which first categorizes theories as Grand, Middle-Range, or Micro level,^[22]^ and then assigns functional attributes, descriptive, explanatory, or predictive, through content analysis of the theoretical propositions.^[23]^ For research question three, a meta-analysis was conducted on self-management behaviors and self-efficacy, using data from the post-intervention time point closest to 3 months (Figures 2-4). This time point was selected a priori because it represents a clinically meaningful window in stroke recovery, sufficiently distant from the immediate post-intervention period to assess the short-term consolidation of self-management skills, yet commonly reported across studies, facilitating data pooling. Standardized mean differences (SMDs) with *95% CIs* were calculated using a random-effects model. Subgroup analyses and meta-regression (e Figures 7-9,13-14; e Table 5) explored effect modifiers and heterogeneity. A separate meta-analysis was performed for self-efficacy theory (Figure 4). Effect sizes were interpreted using Cohen’s conventional benchmarks for the standardized mean difference (SMD): 0.2 represents a small effect, 0.5 a medium effect, and 0.8 a large effect.^[24, 25]^

**Figure 2.**
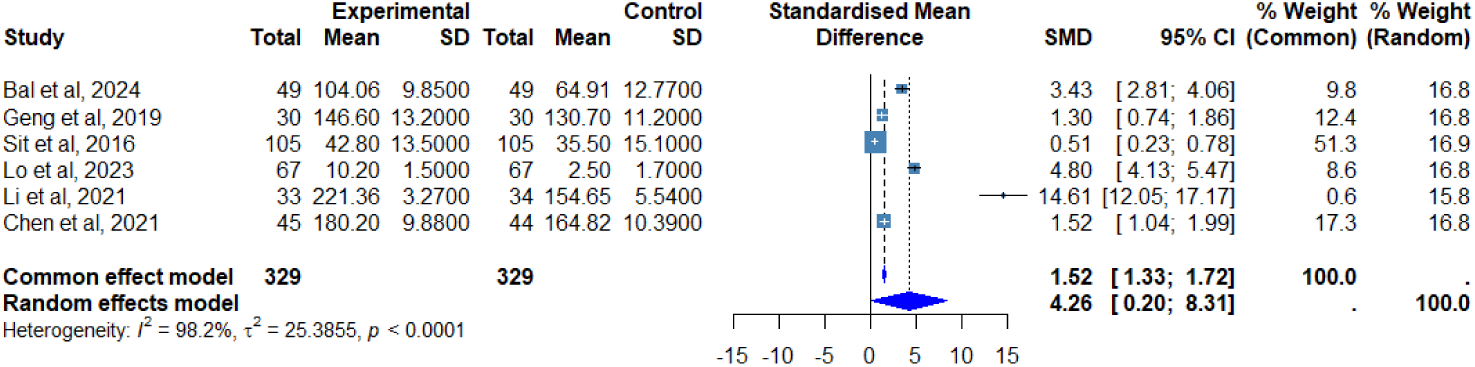
Meta-Analysis for the effect of TMFs-based self-management interventions on self-management behaviors in stroke survivors.

**Figure 3.**
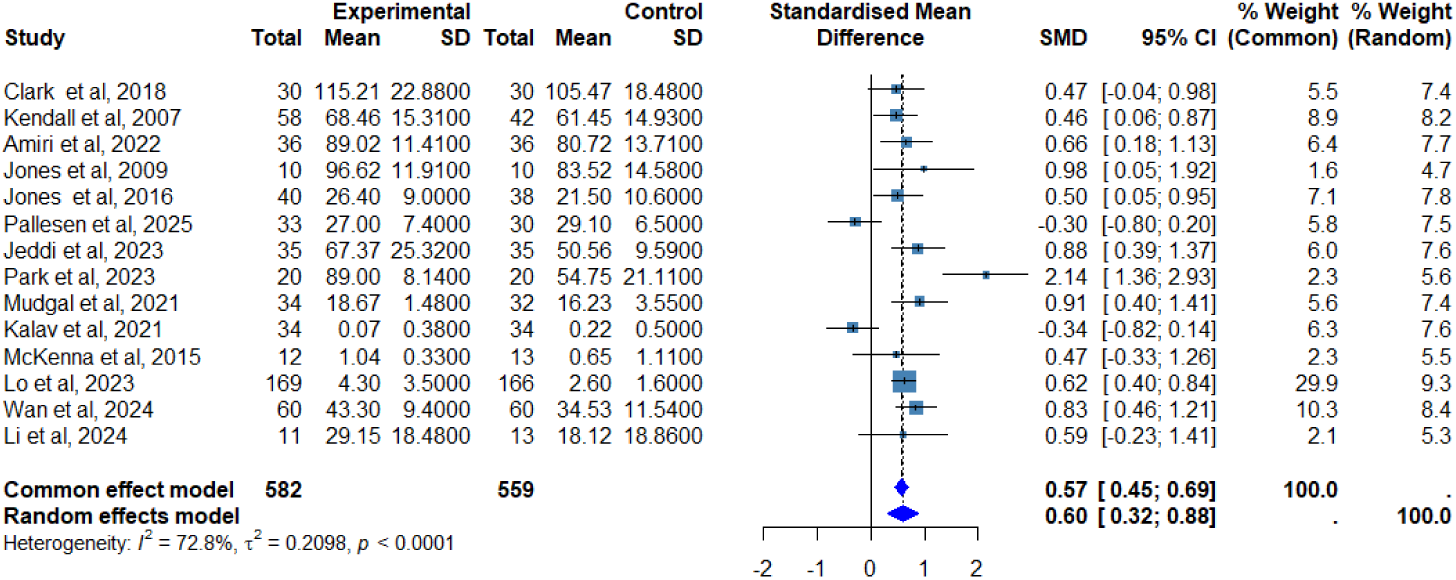
Meta-Analysis for the effect of TMFs-based self-management interventions on self-management self-efficacy in stroke survivors.

**Figure 4.**
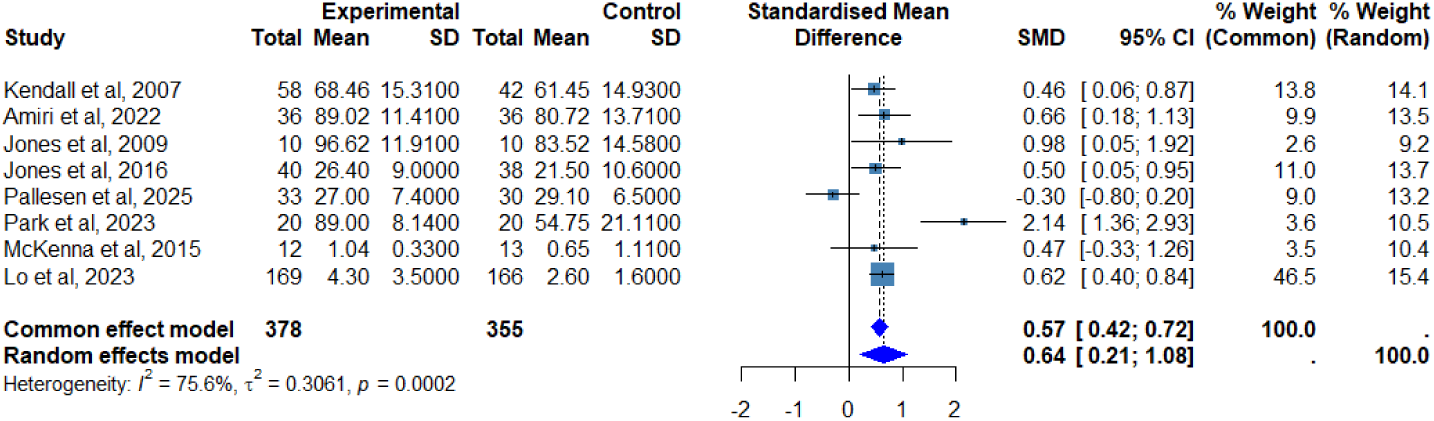
Meta-Analysis for the effect of self-efficacy theory-based self-management interventions on self-management self-efficacy in stroke survivors.

### 2.5 Assessment of Risk of Bias

Two reviewers independently assessed the risk of bias used the Cochrane Risk of Bias Tool 2 checklist^[26]^ and ROBINS-I^[27]^ (e Table 4).

### 2.6 Statistical Analysis

A meta-analysis was conducted only when a minimum of two independent studies provided comparable data for a given outcome. All analyses were performed in R (version 4.5.1) and Stata 17.0 using the metan package. A two-sided *P* value < 0.05 was considered statistically significant unless otherwise stated. We pooled effect sizes as standardized mean differences (*SMD*s) under a random-effects model.^[24]^ Heterogeneity was quantified with the *I²* statistic, with values interpreted as follows: < 30% (minimal), 30–50% (moderate), and > 50% (substantial)^[24]^. Publication bias was assessed both visually through funnel plot inspection (e Figure 1, 4, 10) and statistically using Egger’s regression test (e Table 6)^[28]^. In cases of significant funnel plot asymmetry, the trim-and-fill method (linear estimator, random-effects) was implemented to impute potentially missing studies and recalculate the adjusted pooled effect size under the assumption of a symmetric distribution (e Figures 2, 5, 11)^[29]^. To assess the robustness of the findings, sensitivity analyses were performed, including leave-one-out analysis, re-estimation with alternative random-effects estimators, and identification of statistical outliers (e Figures 3, 6, 12).^[24]^ Subgroup analyses were conducted to explore the influence of categorical moderators and potential sources of heterogeneity based on study design, country, and the type of TMFs (e Figures 7-9, 13-14). Meta-regression was employed to examine the effects of continuous moderators, including mean participant age and intervention duration (e Table 5).

## 3. Results

### 3.1 Study selection and characteristics

Of the 14,234 articles identified from databases and citation searching, 11,495 were excluded after duplicate removal and title/abstract screening. After full-text review of 590 reports, 32 original studies met the inclusion criteria, of which 20 were included in the meta-analysis (Figure 1).

The 32 original studies were published between 2006 and 2024, spanning four continents, with the majority from Mainland China (n=12), 26 studies were randomized controlled trials;^[30–39, 42–50, 52, 54, 56, 58–61]^ five were quasi-experimental^[41, 51, 53, 55, 57]^ and one was a single-group pre-post design.^[40]^ Participants were adults with mild-to-moderate stroke who had been discharged to home or community settings. Sample sizes ranged from 10 to 335. Thirty-one studies utilized control groups that received self-management interventions without TMFs; one study employed a single-group pre-post design.^[40]^

Interventions were delivered via diverse formats (e.g., coaching, workbooks, virtual reality) over four to 24 weeks,^[33, 34, 40]^ which were administered by multidisciplinary teams including nurses,^[38]^ therapists,^[35]^ social workers,^[50]^ and peer volunteers.^[33]^ Core components included personalized goal-setting,^[35, 36, 39]^ disease education,^[43, 46, 54]^ peer support,^[32, 44]^ and self-monitoring.^[55, 58]^ Primary outcomes focused on self-management self-efficacy (most frequently measured with the Stroke Self-Efficacy Questionnaire, SSEQ) and self-management behaviors (e.g., Stroke Self-Management Behaviors Performance Scale, SSMBPS/Stroke Self-Management Behaviors Performance Scale, SSBPS). Secondary outcomes included health-related quality of life, activities of daily living, and emotional status. A detailed summary is provided in Table 1.

### 3.2 Characteristics of TMFs

Across the 32 included interventions, 16 distinct TMFs were identified (e Table 2). All were classified as middle-range,^[62–78]^ offering testable propositions suitable for behavior-change interventions in chronic disease;^[22, 23]^ no grand or micro level theories were used.

Regarding theory type, 14 of 16 TMFs were believed to have explanatory roles, elucidating mechanisms such as how self-efficacy influences behaviors^[32, 35–38, 39–41, 55]^ or how environmental and personal factors interact.^[31, 42, 61]^ Seven also possess predictive functions, positing that changes in specific variables (e.g., self-efficacy, perceived barriers/benefits) lead to predictable outcome. A smaller subset (Timing It Right Framework and Lorig and Holman’s Self-Management Theory) were descriptive, defining stages of behaviors^[69]^ or delineating self-management components.^[72, 73]^

In terms of core concepts, self-efficacy emerged as a central construct across four different TMFs.^[63, 65, 66]^ Most TMFs feature provide clear propositions that map pathways of change (e.g., self-efficacy theory proposes that increased self-efficacy leads to improved self-management behaviors, thereby contributing to better health outcomes).^[65]^ These propositions informed the logic models and outcome evaluations of the included interventions.

Geographically, 15 TMFs originated in the United States and one in Canada.^[69]^

### 3.3 How TMFs guided intervention design

Among the 32 studies, self-efficacy theory was the most frequently adopted framework (13 studies). Interventions leveraged its four core concepts (mastery experience, vicarious learning, verbal persuasion and emotional regulation) to strengthen post-stroke confidence.^[65]^ Examples include workbooks for action-planning and progress tracking,^[35]^ success stories shared in group sessions,^[36]^ graded virtual reality tasks,^[34]^ and nurses’ timely encouragement with next-step goal-setting^[41]^. Six studies combined multiple TMFs to address multi-level barriers,^[35, 42, 45, 47–49]^ for instance, Cadilhac et al. integrated the Information-Motivation-Behavioral Skills (IMB) Model and social cognitive theory to tailore-health goals.^[48]^

Across the included studies, TMFs supported intervention design through four key functions: measurement selection, intervention logic, concept operationalization, and hypothesis testing of variable relationships.

Measurement selection: Self-efficacy theory-based interventions used the Stroke Self-Efficacy Questionnaire (SSEQ);^[28, 33-35 38-41, 55]^ Orem’s self-care theory guided use of the Exercise of Self-Care Agency Scale (ESCA).^[59]^

Intervention logic: The Information-Knowledge-Attitude-Practice (IKAP) Model structured programs into four sequential steps: providing information (I), building knowledge (K), fostering a positive attitude (A), and promoting the adoption of practice (P).^[60]^

Concept operationalization: Lorig and Holman’s Self-Management Theory translated self-management behaviors into measurable items via the Stroke Self-Management Behaviors Performance Scale (SSBPS).^[37]^

Variable-relationship hypotheses: Orem’s self-care theory hypothesis that nursing support could enhanced self-care agency firstly, then improved quality of life.^[43, 44]^

A detailed summary of TMF application is provided in e Table 3.

### 3.4 TMFs-based interventions improve self-management behaviors in stroke survivors

In six studies,^[33, 42, 43, 46, 51, 56]^ 658 participants were included in a meta-analysis (Figure 2). Compared to usual care, TMFs-based self-management interventions showed a substantial and significant effect on self-management behaviors (*SMD* = 4.26, 95% *CI:* 0.20-8.31), with high heterogeneity (*I²* = 98.2%, τ*²* = 25.40, *p* = 0.0001).

Each of these six studies was grounded in a distinct theoretical framework (Orem’s Self-Care Theory, Integrated Behavioral Model, Health Empowerment Theory, Self-efficacy Theory, Theory of Planned Behavior, and Health Promotion Theory). Consequently, a formal subgroup analysis to compare the effectiveness of different TMFs could not be performed due to the single-study-per-theory constraint.

### 3.5 TMFs-based interventions improve self-efficacy in stroke survivors

A total of 14 studies, 1,141 participants were included in a meta-analysis to investigate whether self-management interventions based on TMFs improved their self-management efficacy (Figure 3).^[30, 31, 34–36, 38–41, 45, 49, 52, 54, 57]^ TMFs-based self-management interventions significantly improved self-management efficacy compared with usual care (*SMD* = 0.60, 95%*CI*:0.32-0.88), with moderate to high heterogeneity (*I²* = 72.8%, τ² = 0.21, *p* = 0.0001).

The 14 studies synthesized in this analysis were grounded in a variety of TMFs. Eight of these studies were explicitly based on self-efficacy theory; the remaining six studies each employed different TMFs (The Theory of Modeling/Role-Modeling, Social Cognitive Theory, Health Promotion Theory, Chronic Care Model, Person-Environment-Occupation Performance Model, and Behavioral Activation Theory). Given that only self-efficacy theory was represented by multiple studies (k=8), we were able to perform a separate meta-analysis for this specific theoretical cluster (Figure 4). For the other TMFs, each was tested in only a single study, precluding a meaningful subgroup analysis to compare their relative effectiveness against one another or against self-efficacy theory.

### 3.6 Self-efficacy theory-based interventions improve self-efficacy in stroke survivors

A total of 8 studies (n = 733 participants) were included in a meta-analysis (Figure 4).^[30, 34–36, 38–41]^ Self-efficacy theory-based interventions significantly improved self-management self-efficacy post-intervention (*SMD* = 0.64, 95% *CI*: 0.21-1.08), with moderate to high heterogeneity (*I²* = 75.6%, τ² = 0.31, *p* = 0.0002).

### 3.7 Subgroup analyses and meta-regression

Exploratory subgroup analyses by study design showed no significant effect modification, either across all TMF-based interventions (χ*²* = 1.08, *p* = 0.30) or within the self-efficacy theory subset (χ*²* = 0.09, *p* = 0.76) (e Figures 9, 14). Subgroups based on country and TMF type were also examined (e Figures 7, 8, 13); however, most strata contained only one trial, precluding formal statistical interpretation. These results are presented descriptively in the appendices.

Meta-regression analyses of continuous moderators are summarized in e Table 5. In the self-efficacy theory subgroup, higher participant age was significantly associated with reduced intervention effect on self-management self-efficacy (β = –0.091, 95%*CI*: –0.159 to –0.023, *p* = 0.009), equivalent to a decrease of 0.09 standard units per additional year of age. In contrast, intervention duration was not a significant predictor in any subgroup. Neither age nor duration showed a significant association with outcomes in subgroups based on multiple TMFs.

### 3.8 Risk of bias and publication bias

The methodological quality of the 32 included studies was assessed using the Cochrane Risk of Bias Tool 2 (RoB 2) for the 26 RCTs^[30–39, 42–50, 52, 54, 56, 58–61]^ and the ROBINS-I tool for the 6 non-RCTs (e Table 4).^[39, 41, 51, 53, 55, 57]^ In 26 RCTs, low risk of bias was noted in nine studies,^[32,33,35,44,46,48,50,59,61]^ moderate risk of bias in 15 studies^[30,31,34,36–39,42,43,47,49,52,56,58,60]^ and high risk of bias in two studies.^[45, 54]^ In six non-RCTs, moderate risk of bias in three studies,^[51,53,55]^ and high risk of bias in three studies.^[40,41,57]^

Publication bias was assessed using funnel plots and Egger’s regression test (e Table 6). For the analysis of self-management behaviors based on multiple TMFs: the funnel plot appeared asymmetrical (e Figure 1), the trim-and-fill algorithm imputed two putative missing studies (e Figure 2), and Egger’s regression test confirmed the bias statistically (*p* = 0.01; e Table 6). For the analysis of self-management self-efficacy based on multiple TMFs: the funnel plot appeared roughly asymmetrical (e Figure 4), the trim-and-fill algorithm imputed four putative missing studies (e Figure 5), and Egger’s regression test indicated no statistically significant bias (*p* = 0.72; e Table 6). For the analysis base on self-efficacy theory: the funnel plot appeared symmetrical (e Figure 10), the trim-and-fill algorithm imputed zero putative missing studies (e Figure 11), and Egger’s regression test indicated no statistically significant bias (*p* = 0.66; e Table 6).

The Leave-one-out sensitivity analyses are presented in e Figures 3, 6, and 12. The exclusion of any single study did not alter the direction of the pooled effect size or cause it to lose statistical significance, indicating that the primary findings are robust.

## 4. Discussion

### 4.1 Summary of principal findings

This systema

### 4.2 Basic characteristics of TMFs: fragmentation and the predominance of middle-range theories

We identified 16 distinct TMFs across 32 included studies, a substantially wider variety than reported in prior reviews restricted to specific care settings.^[79, 80]^ This broader range likely reflects our inclusive search strategy, which imposed no restrictions on intervention setting or survivor residence.

All 16 TMFs were classified as middle-range theories; no grand or micro-level theories were identified. This finding indicates that, within the current empirical literature on stroke self-management, middle-range theories are the predominant form of theoretical guidance actually operationalized in intervention research. Grand theories, by virtue of their high abstraction, are rarely translated into testable interventions, while micro-level theories are often embedded within larger frameworks. Middle-range theories, designed to address concrete problems with testable proposition, offer operational advantages that make them particularly feasible for intervention research^[81, 82]^. In other words, the predominance of middle-range theories should not be equated with empirical superiority over grand or micro-level frameworks. This predominance appears to reflect researchers’ practical feasibility preferences, rather than being grounded in rigorous comparative evidence. To address the evidence gap, future research needs to design interventions that directly compare theories across different levels (e.g., contrasting a macro-level Social Ecological Model with a mid-level Social Cognitive Model).

### 4.3 TMFs guidance

The TMFs in the included studies articulated four core pathways for guiding intervention research.

Operationalization of concepts represents the first key pathway through which TMFs guide research. This process converts abstract constructs into operationalized concepts, which is the basis for choosing appropriate measurement tools for the concepts. The theory clarifies the core connotation and boundaries of the concept. For instance, in Bandura’s self – efficacy theory,^[65]^ the concept of “self-efficacy” is defined as “an individual’s subjective judgment and belief regarding their capacity to successfully execute a particular behavior within a specific context or task. That is, it refers to the level of confidence an individual has in applying their existing skills to achieve the desired goal.” Furthermore, establishing conceptual definitions based on the same TMF across different studies ensures conceptual consistency and enables meaningful comparison of findings, thereby maintaining comparability of research outcomes.

Selection of measurement tools constitutes the second critical pathway of TMF guidance. Theoretical frameworks ensure precise alignment between measurement instruments and conceptual constructs. Instruments developed from specific theories, like the Self-Care Agency Scale grounded in Orem’s self-care theory^[43, 44]^, inherently embody their theoretical foundations. Employing such theory-congruent tools ensures content validity while enabling meaningful comparisons across studies sharing the same theoretical framework.

Intervention logic construction forms the third essential pathway for TMF implementation. Theoretical frameworks provide a structured conceptual foundation for developing coherent intervention mechanisms. For instance, in cerebrovascular disease care, the Information-Knowledge-Attitude-Practice (IKAP) model guides intervention design through a progressive logic: collecting patient information to identify needs; implementing diversified health education to transform disease knowledge; providing psychological support to reshape positive attitudes; and conducting skill-training sessions to solidify healthy behaviors.^[60]^ This approach transforms interventions from conceptually unclear processes into theoretically grounded, testable change mechanisms.

Hypothesis of variable relationships establishes the fourth fundamental pathway of theoretical guidance. TMFs provide testable propositions about conceptual relationships that can be directly translated into specific research hypotheses. For example, self-efficacy theory not only proposes a general association between self-efficacy and health outcomes, but specifically predicts that “self-efficacy levels show positive correlations with quality of life in stroke survivors.”^[36]^ This transitions hypothesis development from speculative exploration to theoretically informed prediction, significantly strengthening the study’s explanatory power and theoretical contribution.

### 4.4 Self-management behaviors

The pooled effect size for self-management behaviors (*SMD* = 4.26, *95% CI*: 0.20–8.31) is exceptionally large and accompanied by extreme heterogeneity (*I²* = 98.2%, τ*²* = 25.40, *p* < 0.001). Several considerations warrant careful attention:

First, the estimate is statistically imprecise. The wide confidence interval (spanning 8.11 units) reflects the small number of studies (k = 6) and indicates considerable uncertainty around the true effect. We believed this might be because each of the six studies was grounded in a distinct theoretical framework (Social Cognitive Theory;^[33]^ Health Belief Model;^[42]^ Orem‘s Self-Care Theory;^[43]^ Health Empowerment Theory;^[46]^ Goal Attainment Theory;^[51]^ Theory of Planned Behavior^[56]^).Because each TMF was represented by only one study, we were unable to perform subgroup analyses to compare their relative effectiveness or to statistically isolate the contribution of theoretical variation to the observed heterogeneity. The pooled SMD of 4.26 therefore represents an average of six conceptually heterogeneous interventions.

Second, methodological factors may have inflated the observed effect size. The six studies varied considerably in their operationalization of self-management behaviors, ranging from medication adherence to complex lifestyle modifications, and employed different measurement instruments (e.g., SSBPS;^[33]^ Stroke Self-Management Behavior Rating Scale;^[42]^ Exercise of Self-Care Agency Scale;^[43]^ Exercise of Self-Care Agency Scale;^[46]^ Health Promoting Lifestyle Profile-II;^[51]^ Stroke Self-Management Behaviors Scale^[56]^). Their varying conceptual scope (generic vs. stroke-specific) and response formats further limit the comparability of the pooled estimate. Small sample sizes^[42, 51]^ and incomplete blinding^[51, 56]^ may have additionally contributed to effect inflation.

Despite these substantial considerations, all six studies consistently reported positive effects in favor of TMFs-based interventions. This consistency suggests that explicit theoretical grounding is feasible and potentially beneficial for enhancing self-management behaviors in stroke survivors. However, the current evidence base does not permit conclusions about which theoretical framework is most effective.

In summary, future research should be advanced in two key directions: first, direct comparative trials that directly contrast the effects of different theoretical frameworks within the same study context are needed; Second, in outcome measurement, given the current absence of an internationally accepted core outcome set for stroke self-management behaviors^[83,84]^, future studies should prioritize disease-specific instruments with established evidence of satisfactory reliability and validity (e.g., the Stroke Self-management Behaviors Performance Scale, SSBPS), and actively contribute to the ongoing standardization of measurement tools. In addition, researchers should provide detailed and transparent reporting of how theoretical constructs are translated into specific intervention components, and ensure adequate sample sizes to support robust comparative analyses.

### 4.5 Self-management self-efficacy

Meta-analysis demonstrated that TMFs-based self-management interventions significantly improved self-management self-efficacy compared with usual care (*SMD* = 0.60, *95% CI*: 0.32–0.88), with moderate to high heterogeneity (*I²* = 72.8%, τ*²* = 0.21, *p* = 0.0001). This finding is consistent with a proposition that self-efficacy serves as a critical psychological mechanism for implementing and sustaining self-management behaviors in this population, functioning as a crucial bridge between knowledge, skills, and actual behavior change.^[13,16,63,65,85,86]^

Compared with a previous meta-analysis by Lau et al.,^[80]^ which reported a small effect (*SMD* = 0.27) based on six trials predominantly grounded in social cognitive theory, our analysis included 14 trials spanning seven distinct TMFs and yielded a larger pooled effect (*SMD* = 0.60). Self-efficacy Theory was the most frequently applied framework in our synthesis (eight of 14 studies). To our knowledge, this is the first meta-analysis to specifically isolate and quantify the effect of interventions explicitly grounded in self-efficacy theory on stroke survivors’ self-management self-efficacy. The pooled effect from this theory-specific subset (*SMD* = 0.64, *95% CI*: 0.21–1.08, *I²* = 75.6%, τ*²* = 0.31, *p* = 0.0002) further supports the feasibility and potential benefit of this theoretical approach. However, as noted in Section 4.2, frequency of use should not be conflated with empirical superiority; whether self-efficacy theory is more effective than alternative frameworks remains untested in direct comparative trials.

Two additional findings warrant consideration. First, subgroup analyses indicated that study design (RCT vs. quasi-experiment) did not significantly modify the pooled effect. This suggests that, within the constraints of the available evidence, the selection of a suitable TMF and its rigorous operationalization may be more critical to intervention success than design type per se. Second, meta-regression revealed a significant negative association between mean participant age and the effect size for self-efficacy outcomes among studies employing self-efficacy theory (β = –0.091, *p* = 0.009). This finding should be interpreted with caution. We postulate that the observed association may reflect the combined influence of multiple age-related factors, including differences in cognitive capacity,^[87]^ digital literacy,^[88]^ baseline efficacy,^[89]^ comorbidity burden,^[90]^ intervention dose,^[89]^ and social support,^[91]^ that could collectively moderate intervention response. However, these explanations remain speculative because none of these candidate moderators were directly measured or empirically tested in the included studies.

In summary, future researchers could: embed formal mediation analyses to examine whether changes in self-efficacy serve as a mediating pathway through which theory-based interventions influence longer-term functional and psychosocial outcomes; conduct direct comparative trials that contrast self-efficacy theory with other TMFs within the same study context, to determine whether this frequently used framework confers any relative advantage; and investigate age-related effect modification using individual participant data meta-analysis or trials designed a priori to test age-by-treatment interactions.

### 4.6 Strengths and Limitations

This review has several strengths. It provides a comprehensive synthesis of theory-based self-management interventions for stroke survivors, systematically identifying and classifying 16 distinct TMFs. By quantifying their effects through meta-analysis and separately examining self-management behaviors and self-efficacy, it offers novel insights into both the overall effectiveness of theory-guided approaches and the comparative evidence base across different theoretical frameworks. It also systematically identifies the critical evidence gaps that prevent conclusions about the relative superiority of any single framework. In addition, the theory-specific analysis of self-efficacy theory (k = 8) represents, to our knowledge, the first meta-analytic evaluation of this frequently used framework in the stroke population.

Several limitations also warrant consideration. First, the methodological quality of included studies was variable: most RCTs were at high or unclear risk of bias in domains related to blinding and allocation concealment, which may have inflated effect estimates. Second, substantial heterogeneity was observed for both primary outcomes. While we used random effects models throughout, the small number of studies, particularly for self-management behaviors (k = 6), limited our ability to conduct pre-specified subgroup analyses or meta-regression to fully explore sources of heterogeneity. Third, the theoretical analysis itself was constrained by the primary literature: with the sole exception of self-efficacy theory, most TMFs were examined in only one to three studies, precluding any meaningful comparison of their relative effectiveness. Incomplete reporting of quantitative data (e.g., means, standard deviations) in some studies further restricted the pool of studies eligible for meta-analysis. Fourth, although we assessed publication bias where possible, the small number of studies per outcome reduced the power of these tests, and the possibility of unpublished null findings cannot be excluded. Finally, the restriction to English-language publications may have introduced language bias.

## 5. Conclusions

Theory-based self-management interventions for stroke survivors are associated with significant improvements in self-management behaviors and self-efficacy. However, the exceptionally large effect for behaviors (SMD = 4.26) is likely inflated and should be interpreted with caution. Self-efficacy theory was the most frequently used framework, but frequency does not imply superiority. Most other theories lack sufficient evidence for reliable comparison. Future research could prioritize direct comparative trials and standardized outcome measurement to advance the field.

## Conflict of Interest statement

The Authors declare that there is no conflict of interest.

**PROSPERO registration number:** CRD420251075002.

## Data Availability

All data produced in the present study are available upon reasonable request to the authors

## Supplemental Online Content

**Note:** the figures in this supplement are organized according to the following three outcome domains: (a) the effects of TMFs-based interventions on self-management behaviours, (b) the effects of TMFs-based interventions on self-management self-efficacy, and (c) the effects of self-efficacy theory-based interventions on self-management self-efficacy.

**e Figure 1.**
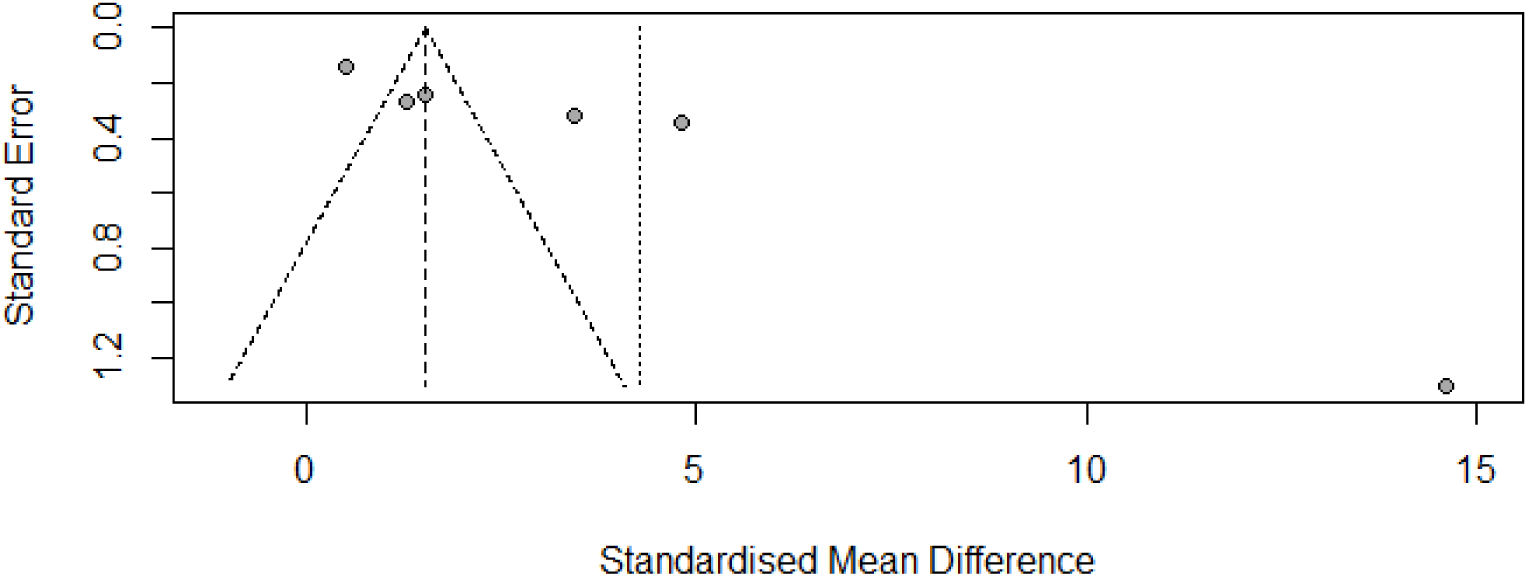
Funnel plot assessing publication bias for TMFs-based interventions on self-management behaviors.

**e Figure 2.**
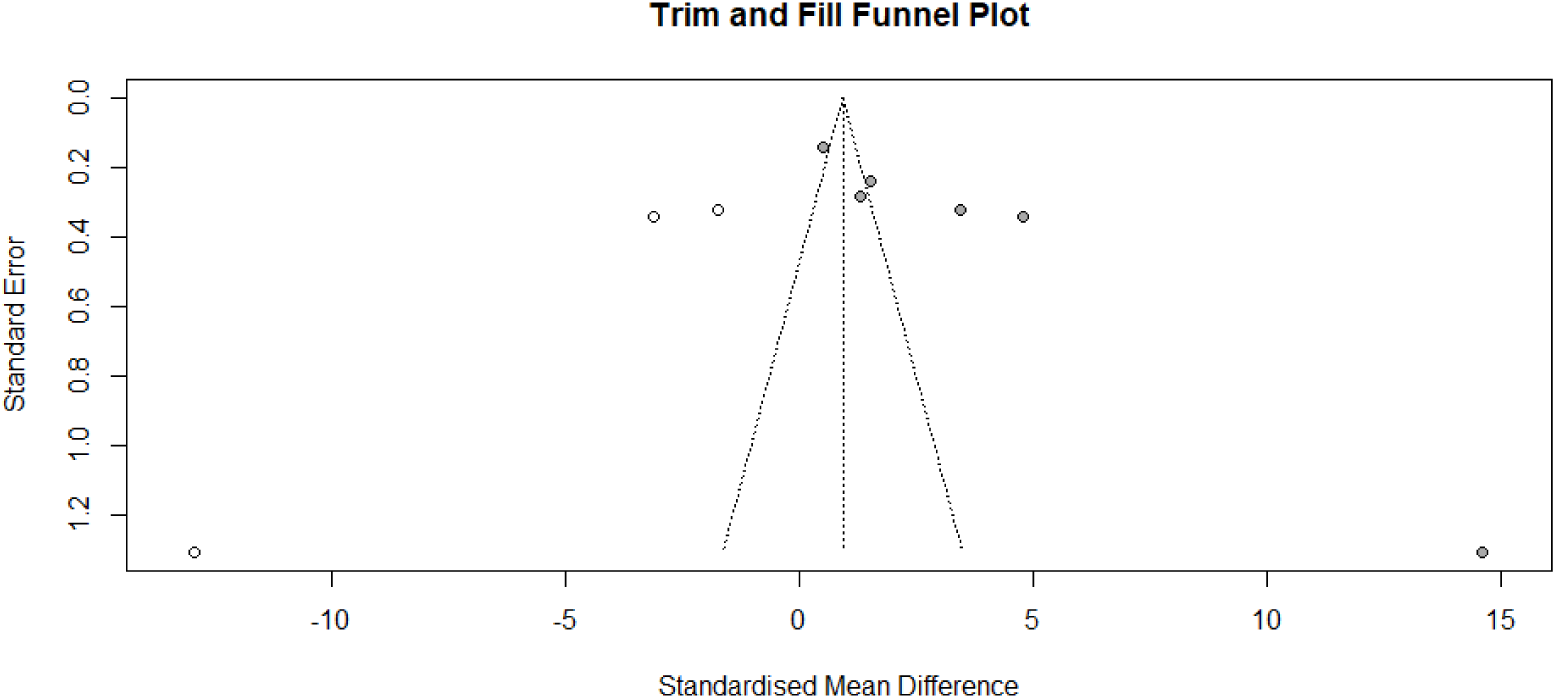
Trim and Fill Funnel Plot assessing publication bias for TMFs-based interventions on self-management behaviors.

**e Figure 3.**
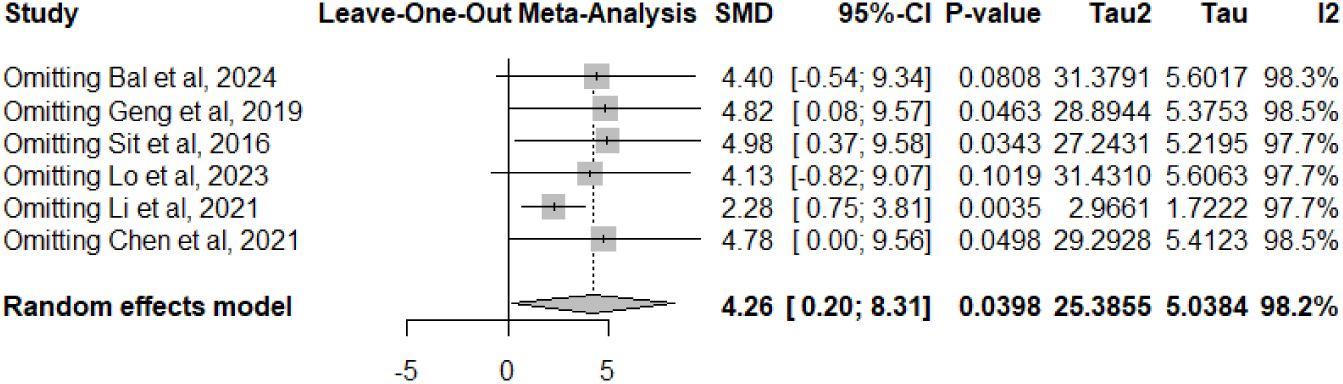
Leave-one-out Meta-analysis for TMFs-based interventions on self-management behaviors.

**e Figure 4.**
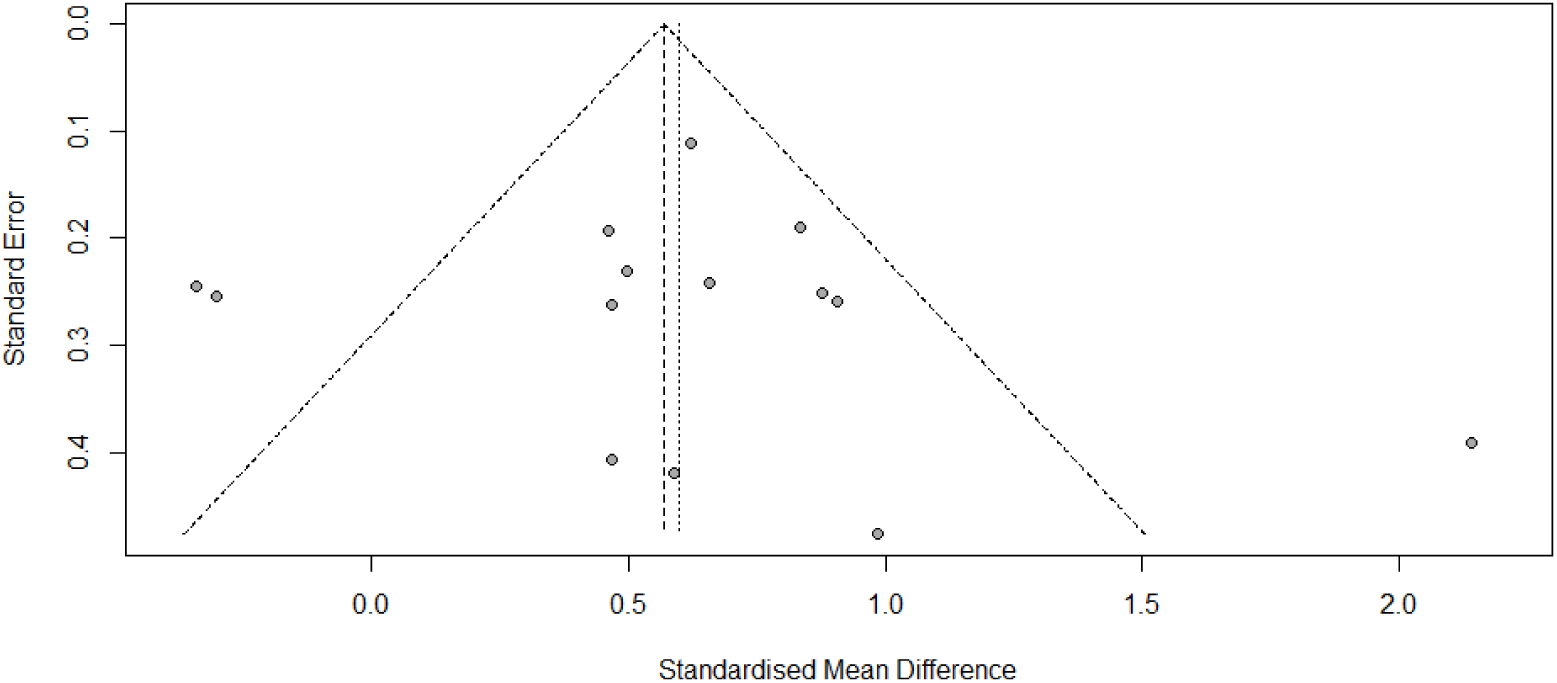
Funnel Plot assessing publication bias for TMFs-based interventions on self-management self-efficacy.

**e Figure 5.**
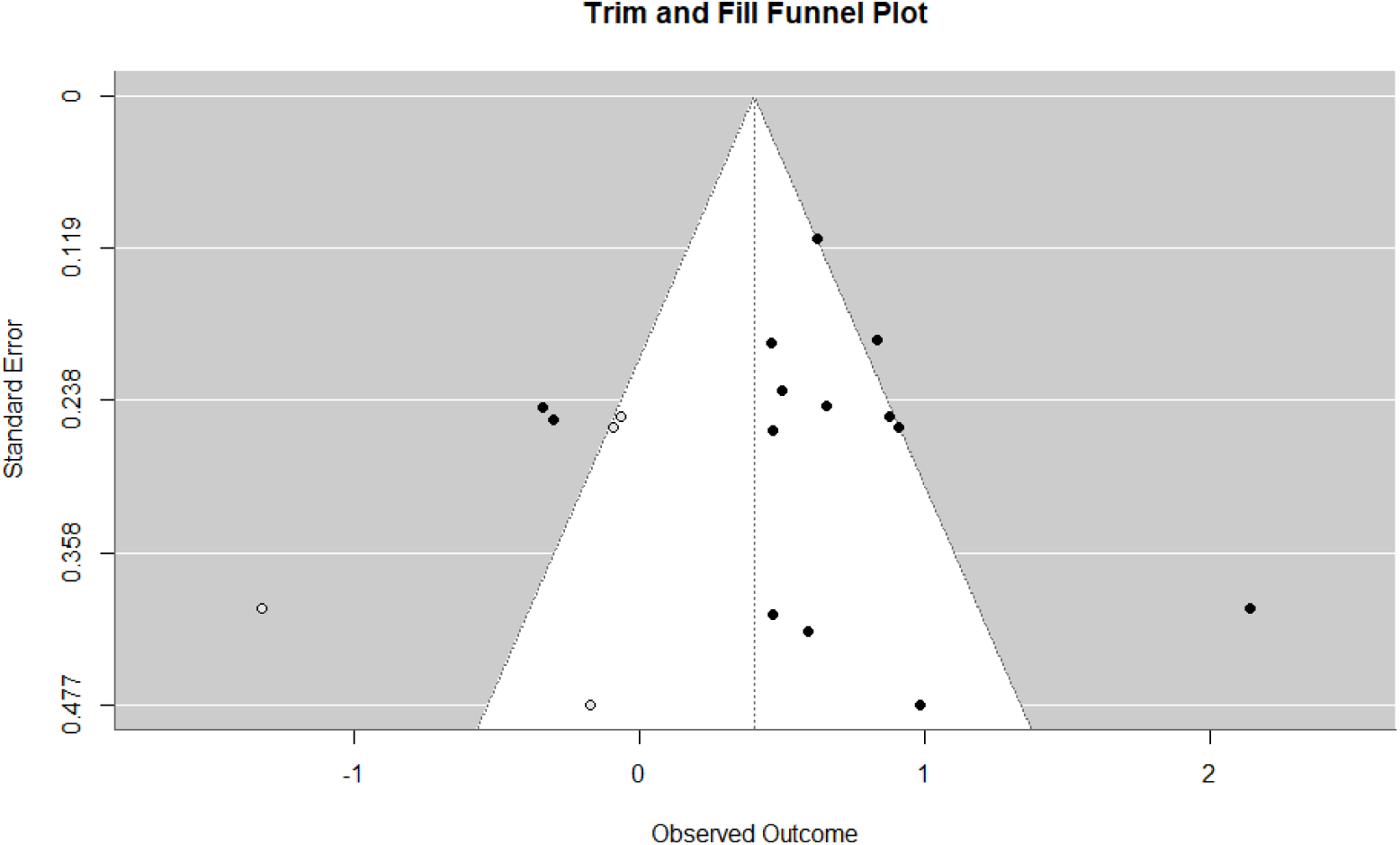
Trim and Fill Funnel Plot assessing publication bias for TMFs-based interventions on self-management self-efficacy.

**e Figure 6.**
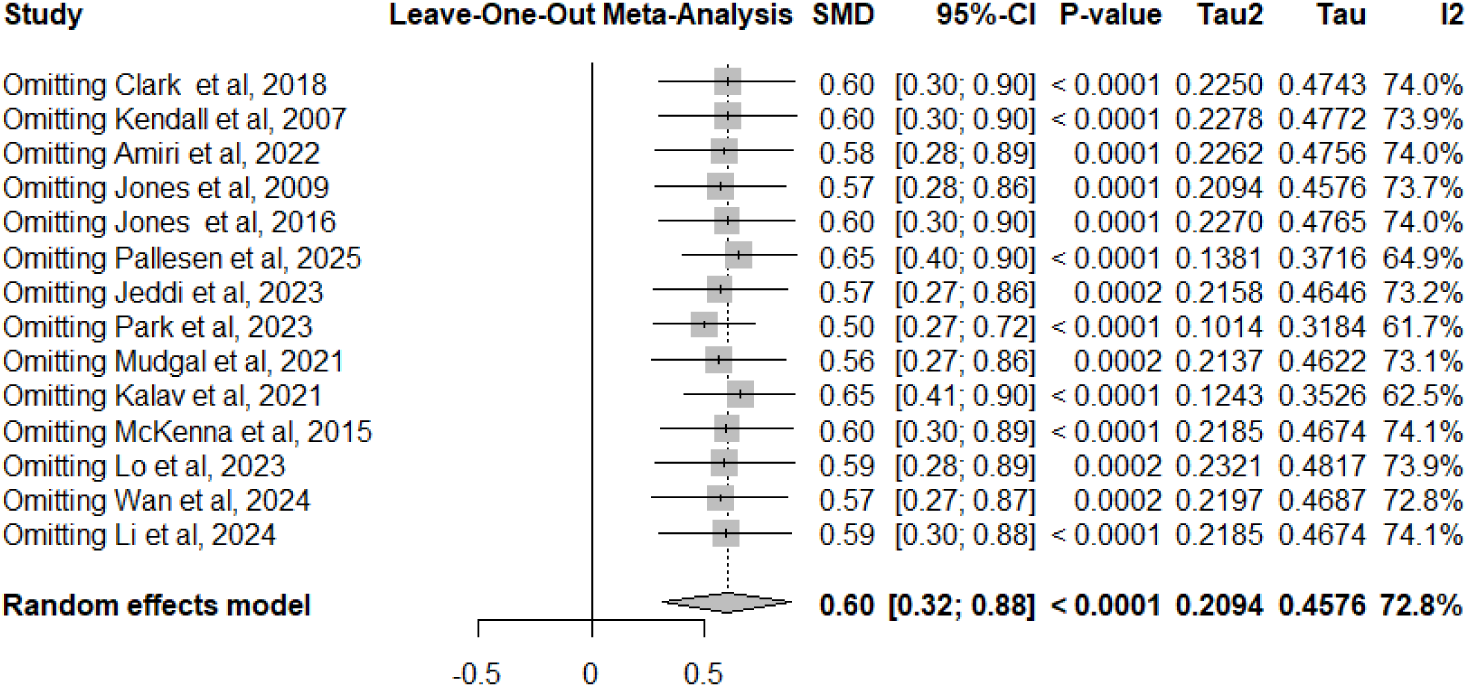
Leave-one-out Meta-analysis for TMFs-based interventions on self-management self-efficacy.

**e Figure 7.**
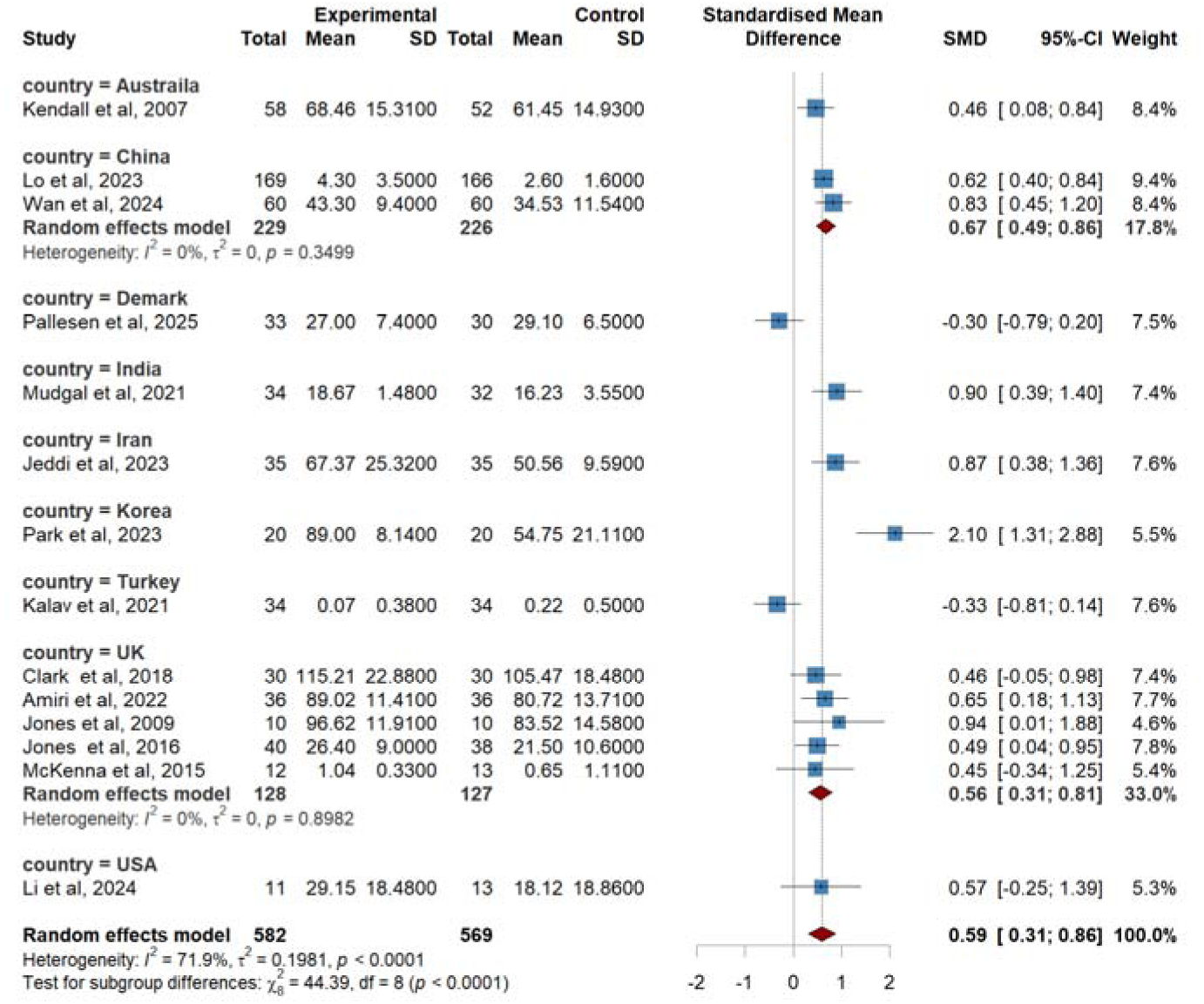
Subgroup analysis by countries for the effects of TMFs-based interventions on self-management self-efficacy.

**e Figure 8.**
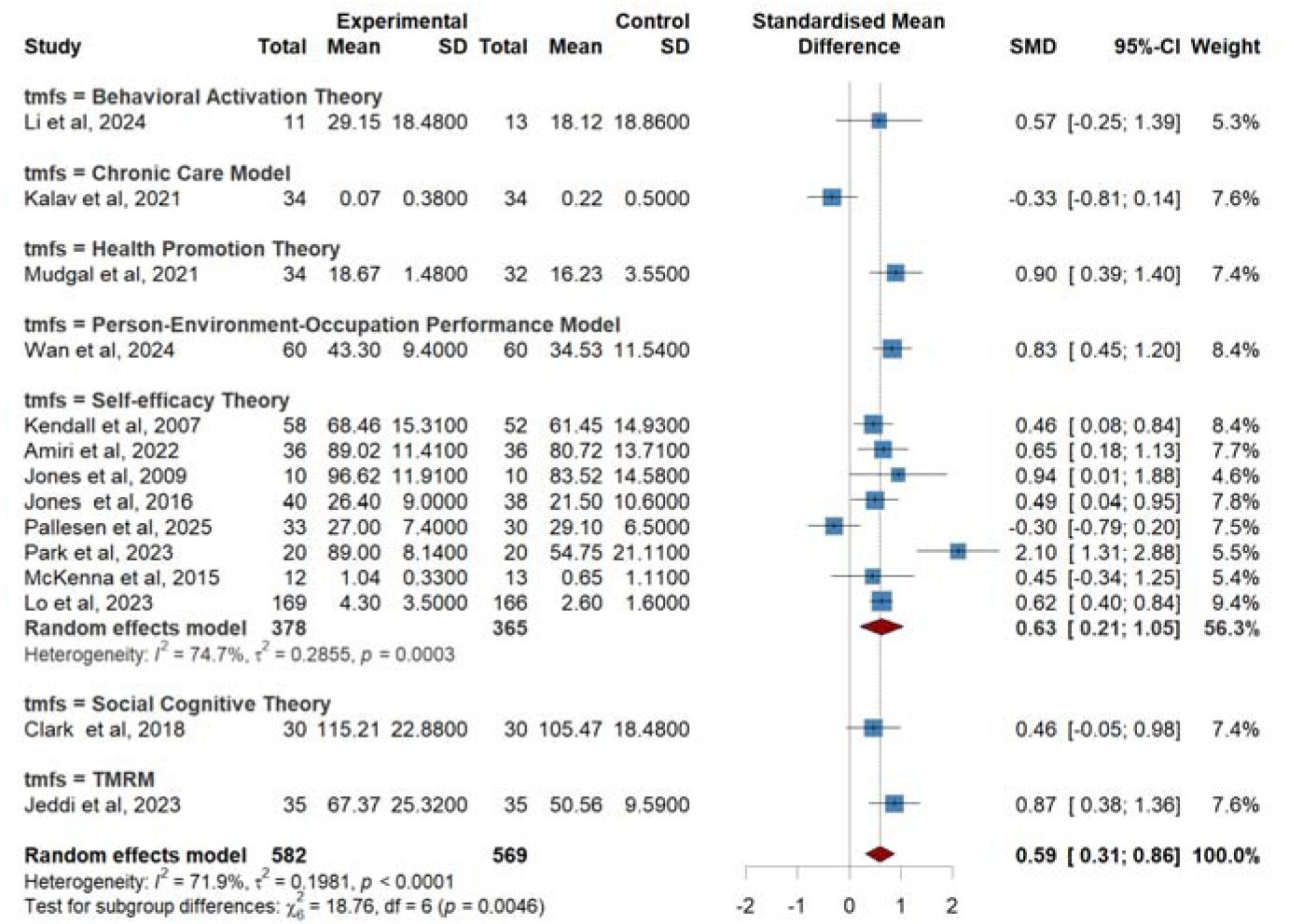
Subgroup analysis by TMFs for the effects of TMFs-based interventions on self-management self-efficacy.

**e Figure 9.**
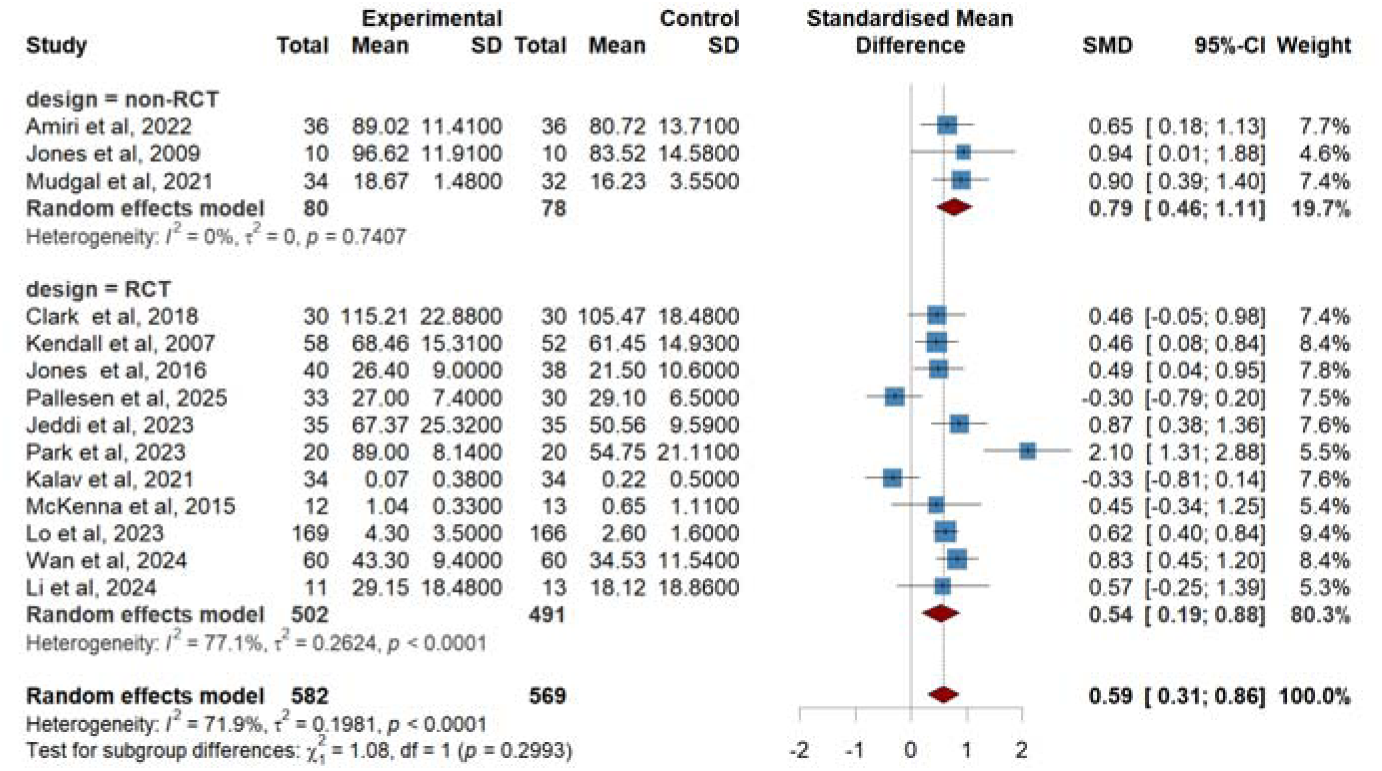
Subgroup analysis by study designs for the effects of TMFs-based interventions on self-management self-efficacy.

**e Figure 10.**
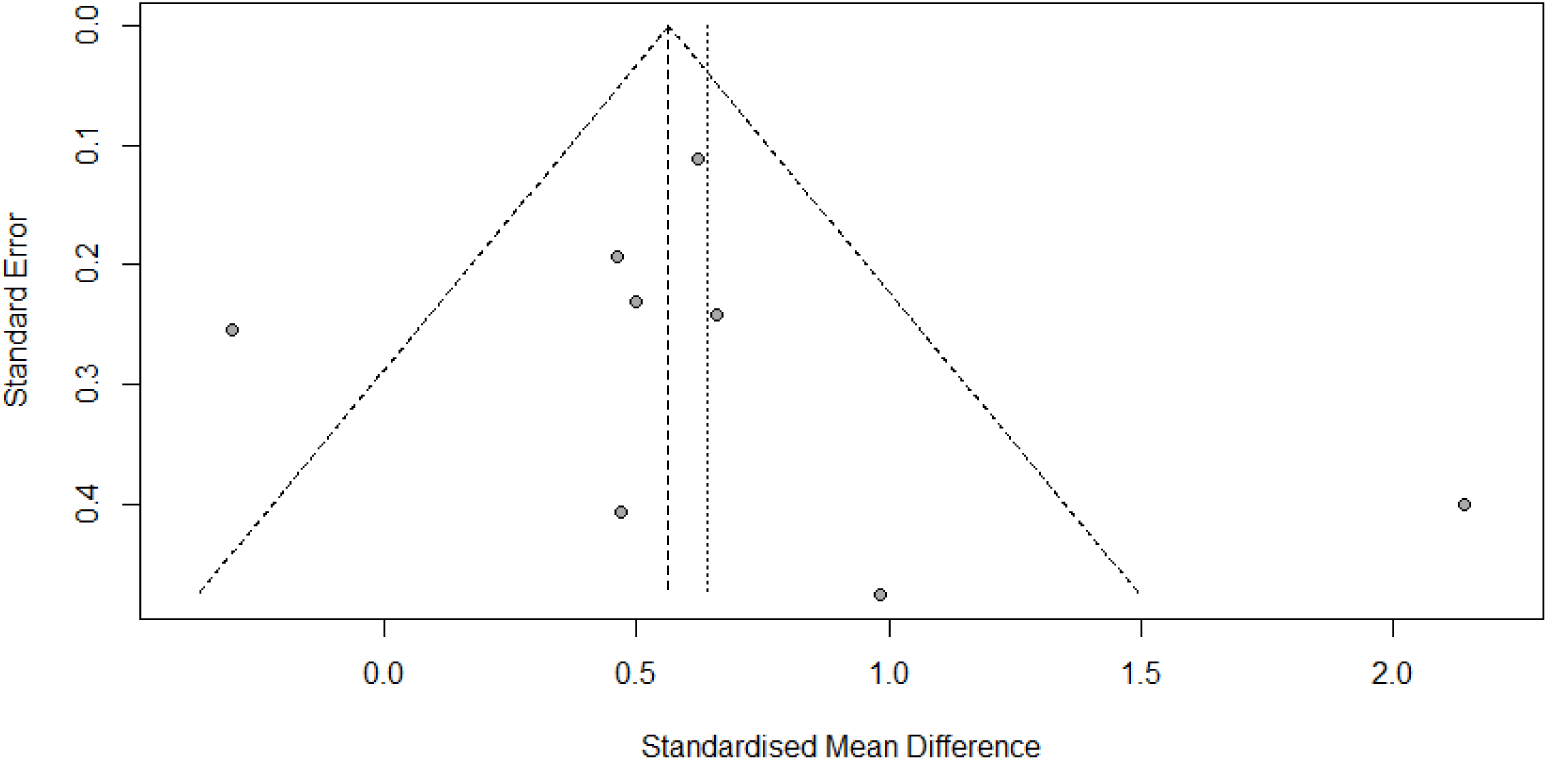
Funnel Plot assessing publication bias for Self-efficacy theory-based interventions on self-management self-efficacy.

**e Figure 11.**
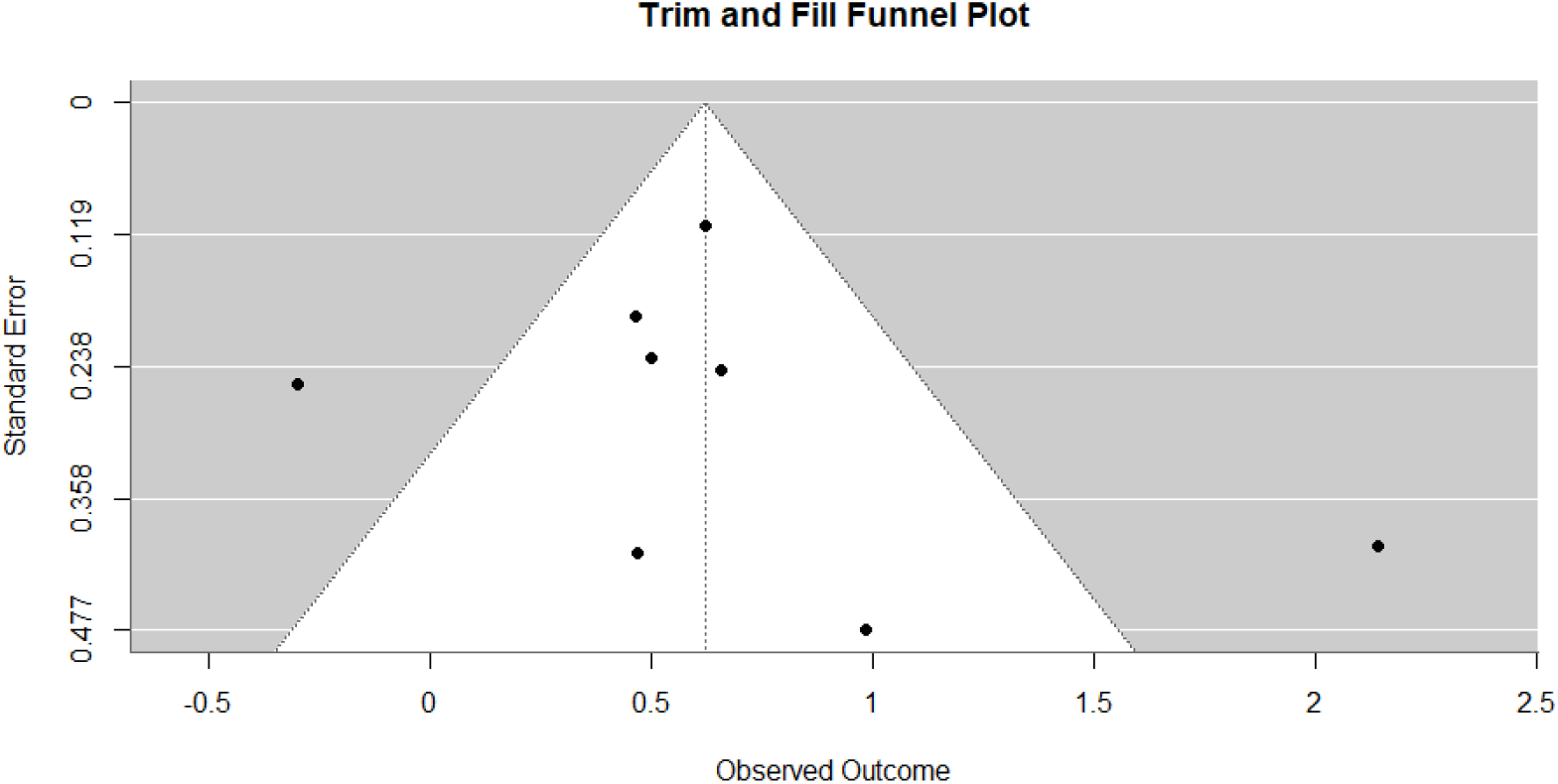
Trim and Fill Funnel Plot assessing publication bias for Self-efficacy theory-based interventions on self-management self-efficacy.

**e Figure 12.**
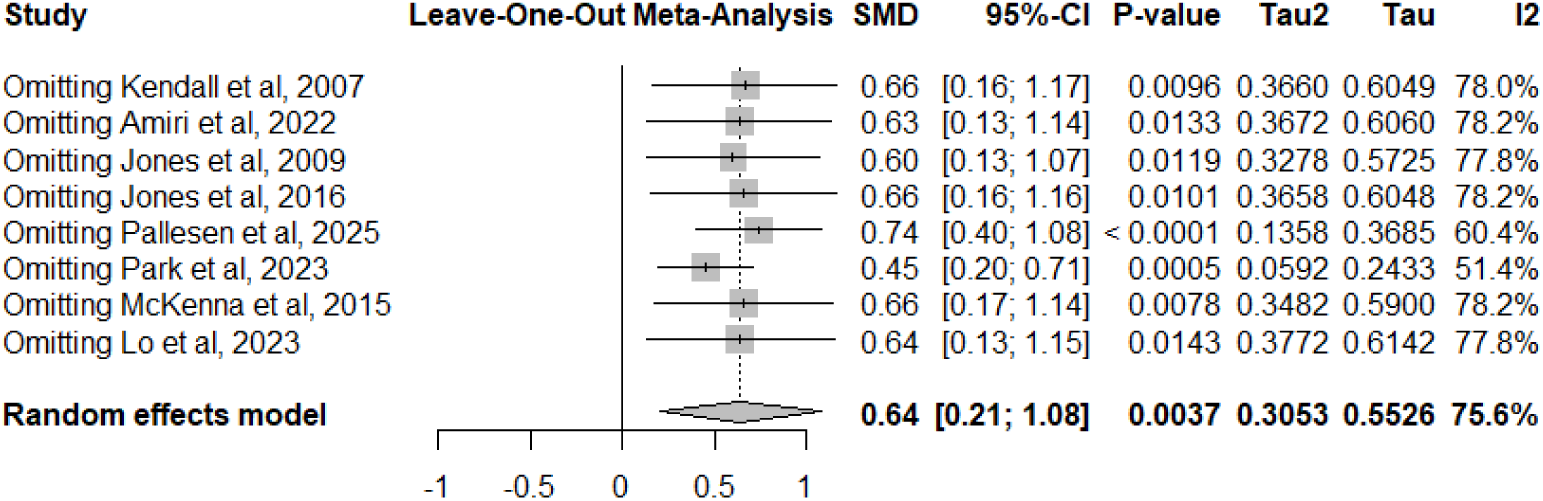
Leave-Out Meta-Analysis for Self-efficacy theory-based interventions on Self-management self-efficacy.

**e Figure 13.**
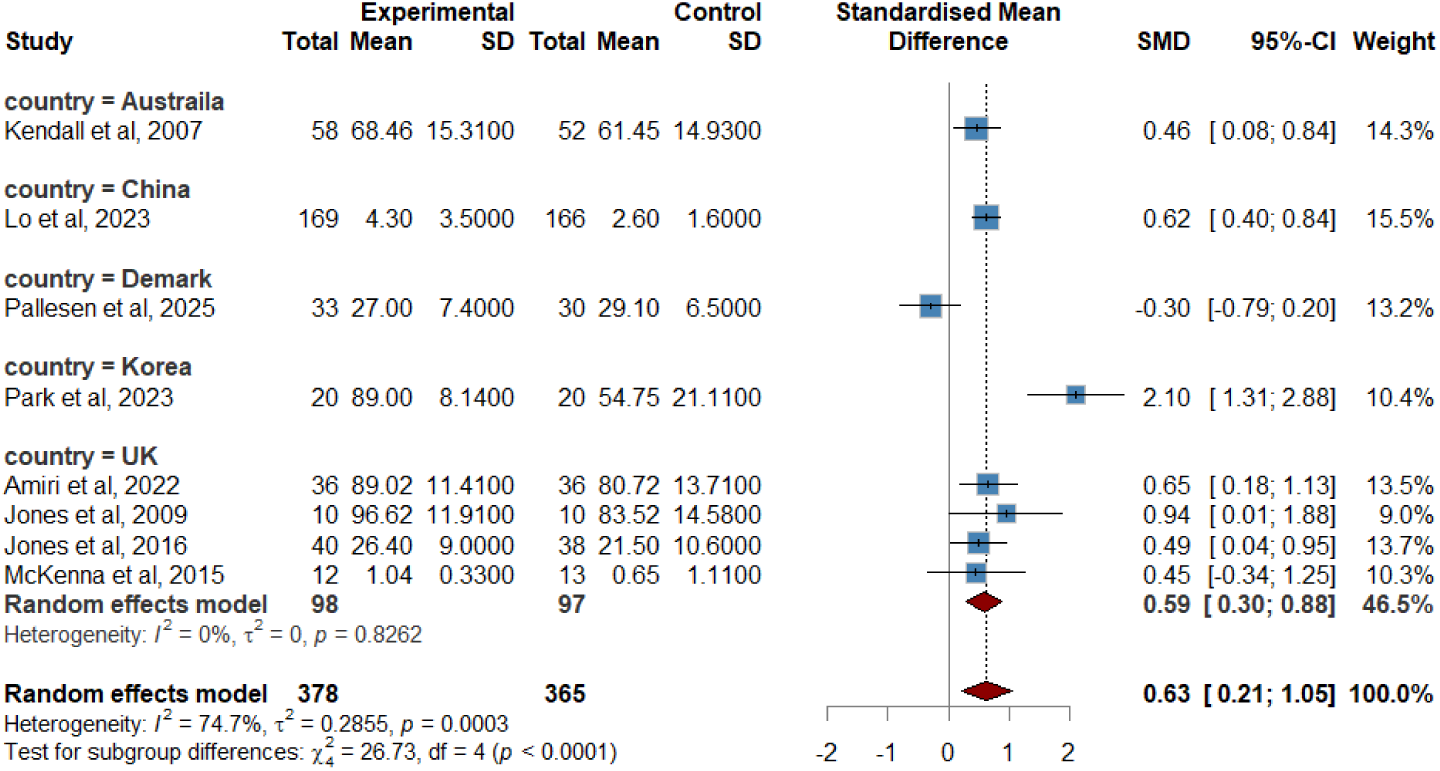
Subgroup Analysis by countries for the effects of self-efficacy theory-based Interventions on self-management self-efficacy.

**e Figure 14.**
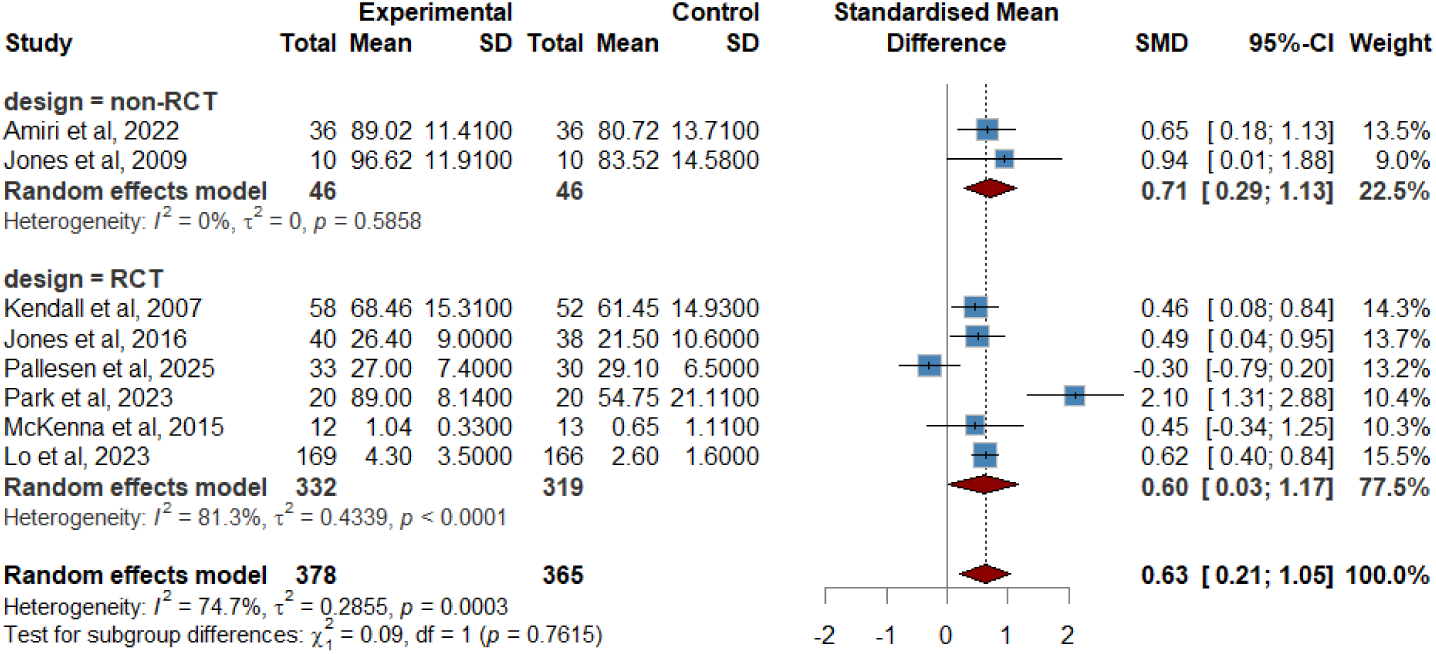
Subgroup Analysis by study designs for the effects of Self-efficacy theory-based Interventions on self-management self-efficacy.

