## Supplementary e Tables 1-6 for "Theory-based self-management interventions for stroke survivors: a systematic review and meta-analysis": Supplementary e Tables 1-6.docx

**Supplementary e Table 1. Search Strategy**

**S1 PubMed**

| 1 | #1 Stroke | (Stroke[MeSH Terms] OR Stroke*[Title/Abstract] OR CVA*[Title/Abstract] OR "Hemiplegia"[Mesh] OR hemipleg*[Title/Abstract] OR hemipar*[Title/Abstract]) OR ((brain*[Title/Abstract] OR cerebrovascular*[Title/Abstract] OR cerebr*[Title/Abstract] OR cerebell*[Title/Abstract] OR intracran*[Title/Abstract] OR intracerebral*[Title/Abstract] OR vertebrobasilar*[Title/Abstract]) AND (diseas*[Title/Abstract] OR accident*[Title/Abstract] OR disorder*[Title/Abstract]) AND (Hemorrhag*[Title/Abstract] OR Ischemi*[Title/Abstract])) |
| --- | --- | --- |
| 2 | #2  Self-management | "Self-Management"[MeSH Terms] OR "Self-Control"[MeSH Terms] OR "Self Care"[Mesh Terms] OR "Self Administration"[Mesh] OR "self restraint*"[Title/Abstract] OR "self control*"[Title/Abstract] OR "self manag*"[Title/Abstract] OR self efficac*[Title/Abstract] OR "self regulat*"[Title/Abstract] OR “self monitor*” [Title/Abstract] |
| 3 | #3 TMF | "theor*"[All Fields] OR "model*"[All Fields] OR "framework*"[All Fields] OR theor*[Title/Abstract] OR model*[Title/Abstract] OR framework*[Title/Abstract] |
| 4 | #4 Intervention | ("Randomized Controlled Trial"[Title/Abstract] OR RCT[Title/Abstract] OR RCTs[Title/Abstract]) OR ((random*[ Title/Abstract] AND (control* OR placebo[Title/Abstract] OR versus[Title/Abstract] OR vs[Title/Abstract] OR group*[ Title/Abstract] OR comparison[Title/Abstract] OR compared[Title/Abstract] OR crossover[Title/Abstract] OR cross-over[Title/Abstract]) AND (trial[Title/Abstract] OR study[Title/Abstract])) OR ((single[Title/Abstract] OR double[Title/Abstract] OR triple[Title/Abstract]) AND (masked[Title/Abstract] OR blind*[ Title/Abstract]))) OR "Clinical Trial"[Title/Abstract] OR intervent*[Title/Abstract] OR treat*[Title/Abstract] OR therap*[Title/Abstract] OR approach*[Title/Abstract] OR program* [Title/Abstract] |
| 5 | #5 Search  Strategy | #1 AND #2 AND (#3 OR #4) |

**S2 Web of Science**

| 1 | #1 Stroke | (TS= (Stroke* OR CVA* OR Hemiplegia OR hemipleg* OR hemipar*)) OR TS= (((cerebrovascular* OR brain* OR cerebr* OR cerebell* OR intracran* OR intracerebral* OR vertebrobasilar*) AND (diseas* OR accident* OR disorder*) AND (Hemorrhag* OR Ischemi*))) |
| --- | --- | --- |
| 2 | #2  Self-management | TS= ("Self Control" OR "Self Care"OR "Self Administration" OR "self restraint*" OR "self manag*" OR "self efficac*" OR "self regulat*" OR "self monitor*") |
| 3 | #3 TMF | TS= ( theor* OR model* OR framework*) |
| 4 | #4 Intervention | (TS= ("Randomized Controlled Trial" OR RCT OR RCTs)) OR (TS= (random* AND (control* OR placebo OR versus OR vs OR group* OR comparison OR compared OR crossover OR cross-over) AND (trial OR study))) OR (TS= ((single OR double OR triple) AND (masked OR blind*))) OR (TS=( "Clinical Trial" OR intervent* OR treat*OR therap* OR approach* OR program*)) |
| 5 | #5 Search  Strategy | #1 AND #2 AND (#3 OR #4) |

**S3 ProQuest Health & Medical Collection**

| 1 | #1 Stroke | (mainsubject(stroke) OR mainsubject(Hemiplegia) OR title(Stroke* OR hemipleg* OR hemipar* OR CVA*) OR abstract(Stroke* OR hemipleg* OR hemipar* OR cva*) OR title((brain* OR cerebr* OR cerebell* OR intracran* OR intracerebral* OR vertebrobasilar* OR cerebrovascular*) AND (diseas* OR accident* OR disorder*) AND (Hemorrhag* OR Ischemi*)) OR abstract((brain* OR cerebr* OR cerebell* OR intracran* OR intracerebral* OR vertebrobasilar* OR cerebrovascular*) AND (diseas* OR accident* OR disorder*) (Hemorrhag* OR Ischemi*))) |
| --- | --- | --- |
| 2 | #2  Self  management | title( "Self Control" OR "Self Care"OR "Self Administration" OR "self restraint*" OR "self manag*" OR "self efficac*" OR "Self Control" OR "Self Car*"OR "Self Administrat*" OR "self restraint*" OR "self manag*" OR "self regulat*" OR "self monitor*") OR abstract( "Self Control" OR "Self Care"OR "Self Administration" OR "self restraint*" OR "self manag*" OR "self efficac*" OR "Self Control" OR "Self Car*"OR "Self Administrat*" OR "self restraint*" OR "self manag*" OR "self regulat*" OR "self monitor*") OR mainsubject(Self-Management) OR mainsubject(Self-Control) OR mainsubject(Self Care) OR mainsubject(Self Administration) |
| 3 | #3 TMF | [(fulltext (theor* OR model* OR framework*) OR abstract (theor* OR model* OR framework*) OR title (theor* OR model* OR framework*))](https://libdb.csu.edu.cn/https/vpn/247/P75YPLURPJYYC7LFPN4C6Z5QNF/myresearch/savedsearches.checkdbssearchlink:rerunsearch/2852960/SavedSearches/$N?site=healthcomplete&t:ac=SavedSearches) |
| 4 | #4  Intervention | abstract (("Randomized Controlled Trial" OR RCT OR RCTs) OR ((random* AND (control* OR placebo OR versus OR vs OR group* OR comparison OR compared OR crossover OR cross-over) AND (trial OR study)) OR ((single OR double OR triple) AND (masked OR blind*))) OR "Clinical Trial" OR intervent* OR treat*OR therap* OR approach* OR program*) OR title(("Randomized Controlled Trial" OR RCT OR RCTs) OR ((random* AND (control* OR placebo OR versus OR vs OR group* OR comparison OR compared OR crossover OR cross-over) AND (trial OR study)) OR ((single OR double OR triple) AND (masked OR blind*))) OR "Clinical Trial" OR intervent* OR treat*OR therap* OR approach* OR program*) |
| 5 | #5 Search  Strategy | #1 AND #2 AND (#3 OR #4) |

**S4 Embase**

| 1 | #1 Stroke | (‘Stroke’ OR ‘Hemiplegia’)/exp OR ((Stroke* OR hemipleg* OR hemipar* OR CVA*) OR ((‘brain*’ OR ‘cerebrovascular*’ OR ‘cerebr*’ OR ‘cerebell*’ OR ‘intracran*’ OR ‘intracerebral*’ OR ‘vertebrobasilar*’) AND (‘diseas*’ OR ‘accident*’ OR ‘disorder*’)) AND (‘hemorrhag*’ OR ‘ischemi*’)): ab,kw,ti |
| --- | --- | --- |
| 2 | #2  Self  management | ('self-management' OR 'self-control' OR 'self care' OR 'self administration')/exp OR ('self care*' OR 'self administrat*' OR 'self restraint*' OR 'self control*' OR 'self manag*' OR 'self efficac*' OR ‘self regulat*’ OR ‘self monitor*’): ab,kw,ti |
| 3 | #3 TMF | (‘theor*’ OR ‘model*’ OR ‘framework*’):ab,kw,ti |
| 4 | #4  Intervention | ((‘Randomized Controlled Trial’ OR ‘RCT’ OR ‘RCTs’) OR ((‘random*’ AND (‘control*’ OR ‘placebo’ OR ‘versus’ OR ‘vs’ OR ‘group*’ OR ‘comparison’ OR ‘compared’ OR ‘crossover’ OR ‘cross-over’) AND (‘trial’ OR ‘study’)) OR ((‘single’ OR ‘double’ OR ‘triple’) AND (‘masked’ OR ‘blind*’))) OR ‘Clinical Trial’ OR ‘intervent*’ OR ‘treat*’ OR ‘therap*’ OR ‘approach*’ OR ‘program*’):ab,kw,ti |
| 5 | #5 Search  Strategy | #1 AND #2 AND (#3 OR #4) |

**S5 The Cochrane Library**

| 1 | #1 Strok | MeSH descriptor (Stroke OR Hemiplegia) OR (Stroke* OR hemipleg* OR hemipar* OR CVA*) OR ((vertebrobasilar* cerebrovascular*) AND (diseas* OR accident* OR disorder*) AND (Hemorrhag* OR Ischemi*)):ti,ab,kw |
| --- | --- | --- |
| 2 | #2  Self  management | MeSH descriptor ((Self Care) OR (Self Management) OR (Self Control) OR (Self Administration)) OR ((Self NEXT Care*) OR (Self NEXT Administrat*) OR (self NEXT restraint*) OR (self NEXT control*) OR (self NEXT manag*) OR (self NEXT efficac*) OR (self NEXT regulat *) OR (self NEXT monitor*)):ti,ab,kw |
| 3 | #3 TMF | (theor* OR model* OR framework*):ti,ab,kw, |
| 4 | #4  Intervention | (("Randomized Controlled Trial" OR RCT OR RCTs) OR (random* AND (control* OR placebo OR versus OR vs OR group* OR comparison OR compared OR crossover OR cross-ove) AND (trial OR study)) OR ((single OR double OR triple) AND (masked OR blind*)) OR "Clinical Trial" OR intervent* OR treat*OR therap* OR approach* OR program*):ti,ab,kw |
| 5 | #5 Search  Strategy | #1 AND #2 AND (#3 OR #4) |

**Supplementary e Table 2. Basic information of the Theories/Models/Frameworks**

| **Basic Information** | | | **Theory Scope** | | | **Theory Type** | | | **Core Concepts** | **Relational Proposition** |
| --- | --- | --- | --- | --- | --- | --- | --- | --- | --- | --- |
| Names | Author  Country | Year | Grand | Middle-Range | Micro | Descriptive | Explanatory | Predictive |  |  |
| The Theory of Modeling/  Role-Modeling (TMRM) ^[62]^ | Erickson HL,  Tomlin E,  Swain MA  USA | 1983 |  | √ |  |  | √ |  | Inherent Worth,  Self-Care Knowledge, Self-Care Resource, Growth Needs, Affiliated-Individuation; Modeling, Role-Modeling, Nurturance. | Modeling → Identify gaps → Role-modeling → Meet needs → Enhance self-care capacity → Health outcomes |
| Social Cognitive Theory ^[63]^ | Albert Bandura  USA |  |  | √ |  |  | √ |  | Triadic Reciprocity,  Observational Learning,  Self-Efficacy, Outcome Expectations,  Reinforcement,  Self-Regulation. | 1. Environment ↔ Personal Cognition ↔ Behavior  2. Observational Learning → Form Outcome Expectations → Enhance self-efficacy → Initiate Self-Regulation → Achieve target behaviors →Reinforcement |
| Chronic Care Model ^[64]^ | Edward Wagner  USA | 1996 |  | √ |  |  | √ |  | Self-Management Support, Delivery System Design, Decision Support, Clinical Information Systems, Organization of Healthcare, Community | Organization of Healthcare → Delivery System Design/ Clinical Information Systems/ Self-Management Support + Community Resource → Health Outcomes |
| Self-efficacy Theory ^[65]^ | Albert Bandura  USA | 1977 |  | √ |  |  | √ | √ | Self-Efficacy, Mastery Experiences, Vicarious Experiences, Verbal Persuasion, Physiological & Affective States, Outcome Expectations. | Mastery Experiences/ Vicarious Experiences/ Verbal Persuasion/ Physiological States → Self-Efficacy↑ → Health Outcomes↑ |
| Health Promotion Model ^[66]^ | Nola Pender  USA | 1982 |  | √ |  |  | √ | √ | **Individual characteristics and experiences:**  Personal Factors,  Prior Behavior  **Behavior-Specific Cognitions:** Perceived benefits/barriers, self-efficacy, activity-related affect, Interpersonal Influence**s,** Situational Influences  **Behavioral Outcomes:** Health-promoting behavior, Commitment to Plan. | Individual characteristics and experience**s** +/ or Behavior-specific cognitions (interaction effects) → Behavior-specific cognitions → Commitment to Plan → Health Outcomes↑ |
| Health Empowerment Theory ^[67, 68]^ | Nelma B. Crawford Shearer  USA | 2009 |  | √ |  |  | √ | √ | **Critical Capacities:**  Cognitive Capacity, Psychological Capacity, Social Capacity  **Empowerment Process:**  Participatory Decision-Making, Resource Accessibility, Self-Management Competence  **Empowerment Outcomes:**  Perceived Control, and Health  Self -determination. | Self-Management Competence →Health Self-Determination↑→Resource Accessibility →Perceived Control↑  ↔ Participatory Decision-Making |
| Timing It Right Framework ^[69]^ | Jill I. Cameron Monique A.M. Gignac  Canada | 2008 |  | √ |  | √ |  |  | Diagnosis, Stabilization, Preparation, Implementation, Adaptation. | NA |
| Information - Knowledge - Attitude - Practice (IKAP) Model ^[60]^ | Mayo  USA | 1950s |  | √ |  |  | √ |  | Information, Knowledge, Attitude, Practice. | Information → Knowledge  → Attitude →Practice |
| Theory of Planned Behavior ^[63]^ | Ajzen  USA | 1985 |  | √ |  |  | √ | √ | Attitude toward Behavior, Subjective Norm, Perceived Behavioral Control (PBC), Behavioral Intention, Actual Behavior, Behavioral Beliefs, Normative Beliefs, Control Beliefs, Actual Control. | Belief-driven → Behavioral intention → Actual behavior↑  Belief-driven:  (1) Behavioral Beliefs → Attitude toward Behavior → Intention →Behavior  (2) Normative Beliefs → Subjective Norm →Intention →Behavior  (3) Control Beliefs → PBC→ Intention →Behavior |
| Health Belief Model ^[63]^ | Hochbaum  USA | 1958 |  | √ |  |  | √ | √ | Perceived Susceptibility,  Perceived Severity, Perceived Benefits, Perceived Barriers,  Cues to Action, Self-Efficacy | (1) Perceived Severity **×**  Perceived Susceptibility → Behavioral motivation → Behavioral trigger  (2) Perceived Benefits/ Self-Efficacy↑ →Behavioral possibility↑→Behavioral trigger  (3) Self-Efficacy↓ →Behavioral possibility↓ →Behavioral trigger |
| Person-Environment-Occupation Performance Model ^[71]^ | Charles Christiansen & Carolyn Baum  USA | 1991 |  | √ |  |  | √ |  | Person,  Environment, Occupation, Performance  (Functional output under the interaction of the three elements). | Person ↔  Environment ↔ Occupation  → Performance |
| Lorig and Holman’s Self-Management Theory ^[72,73]^ | Lorig & Holman  USA | 2003 |  | √ | √ | √ |  |  | Problem Solving, Decision Making, Resource Utilization, Patient-Provider Partnership, Action Planning, Self-Tailoring, Emotional Management. | NA |
| Orem’s  Self-Care Theory ^[74,75]^ | Dorothea E. Orem  USA | 1971 |  | √ |  |  | √ | √ | Self-Care, Self-Care Agency, Self-Care Deficit, Self-Care Needs (1) Universal Needs; (2) Developmental Needs;  (3) Healthy Deviant need, and Nursing Systems  (1) Wholly Compensatory System:  (2) Partially Compensatory System.  (3) Supportive-Educative System. | Effective nursing system → Self-care agency↑ → Self-Care needs → Healthy outcomes |
| Behavioral Activation Theory ^[76]^ | Peter M. Lewinsohn  USA | 1971 |  | √ |  |  | √ | √ | Behavioral Avoidance, Behavioral Activation, Contextual Vulnerability, Reward Learning, Activity-Mood Cycle, Goal Grading. | Goal Grading → Behavioral Activation → Reward Learning →Positive emotion↑ → Negative emotions↓ →Break Activity-Mood Cycle  Activity-Mood Cycle:  Contextual Vulnerability → Behavioral Avoidance ↔ Negative emotions. |
| Information-Motivation-Behavioral Skills Model ^[77]^ | Jeffrey D. Fisher & William A. Fisher  USA | 1992 |  | √ |  |  | √ | √ | Information-Motivation-Behavioral Skills. | Information/ Motivation →Behavioral Skills → Healthy Motivation↑ |
| Integrated Behavioral Model ^[78]^ | Martin Fishbein &  Icek Ajzen  USA | 1990s |  | √ |  |  | √ |  | Attitude, Subjective Norm, Perceived Behavioral Control (PBC), Skills, Intention, Environmental facilitators and barriers, Salience,  Habit, Behavior. | Attitude, Subjective, Norm,  PBC → Behavioral Intention  Behavioral Intention，  Skills, Environmental facilitators and barriers, Salience, Habit → Behavior |

**Supplementary e Table 3. Guidance of TMFs in research**

| **Authors year** | **TMF Name** | **Description of the TMF in the study** | **The guidance of the TMF in the study** | | | |
| --- | --- | --- | --- | --- | --- | --- |
|  |  |  | Operationalization of Concepts | Selection of Measurement Tools | Intervention  Logic  Construction | Hypothesis of Variable Relationships |
| Pallesen et al, 2025 ^[30]^ | Self-Efficacy Theory | The self-management intervention was designed to increase self-efficacy, underpinned by Bandura’s concept of self-efficacy. This intervention focused on promoting growth, development and self-efficacy by facilitating participants’ self-management strategies regarding their everyday activities and social network. | √ | √ | √ | √ |
| Wan et al,  2024 ^[31]^ | Person-Environment-Occupation Performance (PEOP) Model | During the intervention, a situational analysis was made regarding the barriers and facilitators on both personal and environmental levels for proposed activities. |  | √ | √ |  |
| Lo et al,  2018 ^[32]^ | Self-Efficacy Theory | Improvements in self-efficacy and outcome expectation were associated with significant improvement in performance of self-management behaviors Participants set recovery goals with action plans recording in a workbook.  Two DVDs containing videos of survivors’ successful management experiences participants discussed with nurses after viewing.  Strategies included acknowledging successes, reinforcing positive outcomes, and encouraging practice  Group sessions facilitated discussions on emotional challenges | √ | √ | √ | √ |
| Lo et al,  2023 ^[33]^ | Self-Efficacy Theory | It is underpinned by Bandura’s principles of self-efficacy and outcome expectation.  Strategies to enhance self-efficacy and outcome expectations adopted. | √ | √ | √ | √ |
| Park et al,  2023 ^[34]^ | Self-Efficacy Theory | The VR-based cognitive rehabilitation program grounded in self-efficacy theory. | √ | √ | √ | √ |
| Jones et al,  2016 ^[35]^ | Self-Efficacy Theory & Social Cognitive Theory | The Bridges stroke SMP is based on social cognition theory and self-efficacy incorporating a patient-held workbook used by rehabilitation professionals to support self-management skills. | √ | √ | √ |  |
| Kendall et al, 2007 ^[36]^ | Self-Efficacy Theory | The CDSM course emphasizes group interaction and support, and reinforces solution-focused behaviors (e.g. problem solving, goal setting, communication aimed at assisting individuals to actively manage the impact of chronic conditions. | √ | √ | √ |  |
| Markle-Reid et al, 2023 ^[37]^ | Lorig and Holman’s Self-Management Theory | Theoretical underpinnings of Lorig and Holman's self-management theory guided the intervention focusing on building skills in problem-solving, decision-making, and resource utilization. | √ | √ | √ |  |
| Lo et al, 2023 ^[37]^ | Self-Efficacy Theory | The VMSCC intervention was grounded in Bandura’s principles of self-efficacy designed to enhance participants’ confidence in managing their condition. | √ | √ | √ |  |
| McKenna et al, 2015 ^[39]^ | Self-efficacy Theory | Strategies to promote specific behaviors that exemplify the hallmarks of self-management... enabling patients to take control by setting small targets, recording progress and problem solving. | √ | √ | √ |  |
| Jones et al,  2009 ^[40]^ | Self-Efficacy Theory | The workbook incorporated sections to increase mastery, vicarious experience and feedback – key sources of self-efficacy described by Bandura. | √ | √ | √ |  |
| Amiri et al, 2022 ^[41]^ | Self-Efficacy Theory | NA | √ | √ | √ |  |
| Li et al,  2021 ^[42]^ | Health Belief Model (HBM) & Theory of Planned Behavior (TPB) | The HBM ignores the influence of external pressures such as subjective norms, TPB ignores the influence of emotional feelings such as threat and fear.  Since a single theory for behavior change can only analyze behaviors from a single perspective, which may result in flaws in behavior explanation, the combination of complementary theories could be applied to improve the intervention effect.  The HBM resolves disease threat cognition, while the TPB exerts social normative effects. |  |  | √ | √ |
| Bal et al,  2024 ^[43]^ | Orem’s  Self-Care Theory | In stroke rehabilitation, supportive-educative nursing system is applied to enhance patients' self-care abilities.  The role of nursing is to provide support through five actions: acting or doing for the patient, guiding the patient, supporting the patient physically/psychologically, teaching the patient, and creating a supportive environment. By enhancing self-care agency, nurses empower stroke patients to independently manage their health deviations, thereby improving quality of life. | √ | √ | √ | √ |
| He et al,  2020 ^[44]^ | Orem’s  Self-Care Theory | As a nursing intervention model developed based on Orem’s Self-Care Theory, collaborative care has a core to give full play to the assisting role of patients’ families and communities to jointly provide health assistance to patients with chronic diseases. | √ | √ | √ |  |
| Li et al,  2024 ^[45]^ | Behavioral Activation Theory (BAT) & Social Cognitive Theory & PEOP Model | ISMART integrates behavioral activation for goal setting, social cognitive theory for skill-building, and the PEOP model to address environmental barriers.  The psychoeducation component of iSMART combines two theoretical approaches: (1) a chronic disease self-management program built on social cognitive or learning theory and (2) community participation and environment management built on the person-environment-occupation-performance model.  The coach worked with each participant to explore their valued life areas and activities and identify treatment goals. derived from the behavioral activation treatment manual. |  |  | √ | √ |
| Sit et al,  2016 ^[46]^ | Health Empowerment Theory | The empowerment process of HEISS enabled participants to set personal goals and develop problem-solving ability congruent with improvements in self-efficacy, participants minimized negative impacts on functional outcomes. |  | √ | √ | √ |
| Damush et al, 2011 ^[47]^ | Social Cognitive Theory & Self-Efficacy Theory | We based our self-management intervention on the Stanford University Chronic Disease Self-Management Program, a program centered around enhancing patient self-efficacy to manage symptoms. | √ | √ | √ | √ |
| Cadilhac et al, 2020 ^[48]^ | Information-Motivation-Behavioral Skills Model (IMB)  & Social Cognitive Theory (SCT) | (1) Information- Motivation-Behavioral Skills Model (IMB)：  Information: Messages were created... under five main categories: administration and general motivation; secondary prevention; health/body function, activities and participation. and environment.  Motivation: Person-centred goals were set with facilitation by a clinician. Participants received daily support messages matched to their personal goals.  Behavioral Skills: Self-management (measured with heiQ) covers skill acquisition to cope with symptoms.  (2) Social Cognitive Theory (SCT):  Facilitating self-efficacy may influence effort invested in health goals and resilience.  A clinician-researcher helped participants set 2 - 3 person-centred goals using a standardized template. |  | √ |  | √ |
| Clark et al, 2018 ^[49]^ | Social Cognitive Theory & Self-Efficacy Theory | The group self-management intervention (adapting the "Bridges" program) is explicitly grounded in Social Cognitive Theory (SCT) and Self-Efficacy Theory (primarily derived from Bandura's work). | √ | √ | √ | √ |
| Reeves et al,2019 ^[50]^ | Chronic Care Model (CCM) | The theoretical underpinnings of the MISTT interventions were guided by core principles of SWCM which were integrated with 3 components of the Chronic Care Model (CCM), specifically: Self-management support, Community resources, and Decision support. | √ | √ | √ | √ |
| Geng et al,  2019 ^[51]^ | Integrated Behavioral Model (IBM) | This study employed the Integrated Behavioral Model (IBM) to guide the TC intervention. The IBM is an extended theory from the Theory of Planned Behavior (TPB). The TPB states: behavioral intention is the determinant of behavioral implementation; and intention is influenced by attitude..., subjective norm..., and perceived behavioral control. IBM adds four constructs: environmental facilitators and barriers, skills to implement the behavior, salience of the behavior, and habit. | √ | √ | √ | √ |
| Jeddi et al, 2023 ^[52]^ | The Theory of Modeling/  Role-Modeling (TMRM) | Clearly describe the three core concepts of TMRM: Self-care Knowledge (SCK), Self-care Resources (SCR) and Self-care Activities (SCA).  Emphasizing the role of nurses: nurses do not work as a self-care agent for the client, but they try to facilitate and nurture self-care resources and knowledge.  Cite five nursing goals: building trust, promoting positive attitudes, promoting control, affirming strengths, and setting mutual health-directed goals. |  | √ | √ |  |
| Jeong et al,  2024 ^[53]^ | Social Cognitive Theory (SCT) | The invention was based on SCT by Bandura and the  ADDIE (Analysis, Design, Development, Implementation,  and Evaluation) model for stroke survivors. The SCT  consisted of three factors: cognitive (self-efficacy and knowledge), behavioral (behavioral skills, intentions, reinforcement), and environmental (observational learning and social support) to improve or change stroke patients' behaviors. |  | √ | √ |  |
| Kalav et al, 2022 ^[54]^ | Chronic Care Model (CCM) | The intervention group received a 12-week StrokeCARE intervention protocol based on the four components of the CCM.: Delivery System Design, Self-Management Support, Decision Support and Decision Support. |  | √ |  | √ |
| Messina et al, 2020 ^[55]^ | Self-efficacy Theory | Self-efficacy can be enhanced through skills mastery, modeling, reinterpretation and social persuasion.  Enhancing self-management and self-efficacy can support individuals and improve their quality of life. | √ | √ | √ | √ |
| Chen et al, 2021 ^[56]^ | Health Promotion Model (HPM) | The goal-oriented self-management intervention was developed based on Pender's health promotion model  The goals and content of the intervention focused on these six dimensions: physical activity, nutrition, stress management, health responsibility, interpersonal support, and self-actualization. | √ |  | √ | √ |
| Mudgal et al,  2021 ^[57]^ | Health Promotion Model (HPM) | Health promotion models like Pender’s Health Promotion Model (HPM) is a beneficial framework. The investigators planned to carry out a study to ascertain the effectiveness of a health promotion model-based visual learning module (HPM-VLM). | √ | √ | √ |  |
| Chen et al, 2018 ^[58]^ | Health Empowerment Theory | The interventions were developed by adopting the key elements of the health empowerment model: facilitating awareness of self-efficacy, knowledge, self-care skills, and building supportive networks. | √ | √ | √ |  |
| Jiang et al,  2021 ^[59]^ | Timing It Right (TIR) Framework | The characteristics of patients' condition in each period were fully mastered considering the needs of emotion, information, evaluation, tools. It can help patients adapt to family/social environments, improve caregivers' nursing ability and patients' adherence, accelerate recovery, providing research opportunities for continuous nursing of chronic diseases. |  | √ | √ | √ |
| Huo et al, 2022 ^[60]^ | Information - Knowledge - Attitude - Practice (IKAP) Model | Improve knowledge → boost self-management ability → change mindset →motivate exercise.  Requires individualized education with focus on communication among staff, families, and patients.  Lays foundation by collecting information (I), then imparting knowledge (K), changing attitude (A), and finally conducting rehabilitation training (P).  Formulate individualized measures based on patient's unique circumstances. | √ | √ | √ | √ |
| Salajegheh et al, 2024 ^[61]^ | Theory of Planned Behavior (TPB) | Behavioral intentions are the primary determinant of behavior, affected by three constructs: attitudes, subjective norms, and perceived behavioral control.  Individuals systematically utilize information and evaluate consequences before engaging in a behavior. With full intent, an individual tends to perform that behavior. | √ | √ | √ | √ |

**Supplementary e Table 4. Quality Assessment of Included Studies Using the Cochrane Risk-of-Bias Tool 2 and ROBINS-I**

**S1. Quality of included studies for RCT using RoB 2.0 (n = 26)**

| **Study** | **Randomization process** | **Deviations from intended interventions** | **Missing outcome data** | **Measurement of the outcome** | **Selection of reported result** | **Total** |
| --- | --- | --- | --- | --- | --- | --- |
| Pallesen et al, 2024 ^[30]^ | Low | Low | Mod | Mod | Low | Mod |
| Wan et al, 2024 ^[31]^ | Low | Low | Low | Mod | Low | Mod |
| Lo et al, 2018 ^[32]^ | Low | Low | Low | Low | Low | Low |
| Lo et al, 2023 ^[33]^ | Low | Low | Low | Low | Low | Low |
| Park et al, 2023 ^[34]^ | Low | Low | Low | Mod | Low | Mod |
| Jones et al, 2016 ^[35]^ | Low | Low | Low | Low | Low | Low |
| Kendall et al, 2007 ^[36]^ | Low | Low | Low | Mod | Low | Mod |
| Markle-Reid et al, 2023 ^[37]^ | Low | Low | Low | Mod | Low | Mod |
| Lo et al, 2023 ^[38]^ | Low | Mod | Low | Mod | Low | Mod |
| McKenna et al, 2015 ^[39]^ | Low | Low | Low | Mod | Low | Mod |
| Li et al, 2021 ^[42]^ | Mod | Low | low | Mod | Low | Mod |
| Bal et al, 2024 ^[43]^ | Low | Low | Low | Mod | Low | Mod |
| He et al, 2020 ^[44]^ | Low | Low | Low | Low | Low | Low |
| Li et al, 2024 ^[45]^ | Low | Mod | High | High | Low | High |
| Sit et al, 2016 ^[46]^ | Low | Low | Low | Low | Low | Low |
| Damush et al, 2011 ^[47]^ | Low | Mod | Low | Mod | Low | Mod |
| Cadilhac et al, 2020 ^[48]^ | Low | Low | Low | Low | Low | Low |
| Clark et al, 2018 ^[49]^ | Low | Mod | Mod | Mod | Low | Mod |
| Reeves et al, 2019 ^[50]^ | Low | Low | Low | Low | Low | Low |
| Jeddi et al, 2023 ^[52]^ | Mod | Low | Mod | Mod | Low | Mod |
| Kalav et al, 2022 ^[54]^ | Mod | Low | Low | High | Low | High |
| Chen et al, 2021 ^[56]^ | Low | Mod | Low | Mod | Low | Mod |
| Chen et al, 2018 ^[58]^ | Mod | Low | Low | Mod | Low | Mod |
| Jiang et al, 2021 ^[59]^ | Low | Low | Low | Low | Low | Low |
| Huo et al, 2022 ^[60]^ | Low | Mod | Low | Mod | Low | Mod |
| Salajegheh et al, 2024 ^[61]^ | Low | Low | Low | Low | Low | Low |

**Note:** Low=Low risk; Mod=Moderate risk; High=High risk

**S2. Quality of included studies for non-RCT using ROBINS-I (n = 6)**

| **Study** | **Confoun-ding** | **Selection of participants** | **Classification of interventions** | **Deviations from intended interventions** | **Missing data** | **Measurement of outcomes** | **Selection of reported result** | **Total** |
| --- | --- | --- | --- | --- | --- | --- | --- | --- |
| Jones et al, 2009 ^[40]^ | Mod | Mod | Low | Low | Low | High | Low | High |
| Amiri et al, 2022 ^[41]^ | High | Mod | Low | Low | Low | High | Low | High |
| Geng et al, 2019 ^[51]^ | Mod | Mod | Low | Low | Mod | Mod | Low | Mod |
| Jeong et al, 2024 ^[53]^ | Mod | Mod | Low | Low | Mod | Mod | Low | Mod |
| Messina et al, 2020 ^[55]^ | Mod | Mod | Low | Low | Mod | Mod | Low | Mod |
| Mudgal et al, 2021 ^[57]^ | High | Mod | Low | Low | Low | Mod | Low | High |

**Note:** Low=Low risk; Mod=Moderate risk; High=High risk

**Supplementary e Table5. Assessment of the Influence of Predictors on Intervention Effectiveness Using Meta-Regression**

| **Subgroup analysis** | **Outcomes** | **Predictor** | **Coefficient (β)** | **95% CI** | ***p* value** | **I^2^** | **R²** | **Studies** |
| --- | --- | --- | --- | --- | --- | --- | --- | --- |
| **Multiple TMFs** | Self-management behaviors | Age | -0.488 | –1.143 to 0.167 | 0.144 | 99% | 17.7% | 6 |
|  |  | Intervention duration | 0.353 | –1.086 to 1.791 | 0.631 | 100% | 0.0% |  |
| **Multiple TMFs** | Self-management self-efficacy | Age | -0.027 | –0.082 to 0.028 | 0.330 | 79% | 0.0% | 14 |
|  |  | Intervention duration | -0.031 | –0.104 to 0.042 | 0.409 | 80% | 0.0% |  |
| **Self-efficacy**  **theory** | Self-management self-efficacy | Age | -0.091 | –0.159 to -0.023 | 0.009 | 64% | 62.9% | 8 |
|  |  | Intervention duration | -0.090 | –0.278 to 0.099 | 0.352 | 85% | 1.1% |  |

**Note:** (1) I² (residual): Represents the percentage of residual heterogeneity that remains unexplained by the predictor variable in the meta-regression model. A higher value indicates that substantial variability in effect sizes persists after accounting for the covariate.
(2) R² (explained): Represents the proportion of between-study variance (τ²) explained by the predictor variable. It indicates how much of the original heterogeneity is accounted for by the covariate (e.g., age or intervention duration).

**Supplementary e Table 6. Assessment of Publication Bias of Egger's Regression Test**

| **Analysis Subgroup** | **Studies** | **Intercept** | **SE** | **95% CI** | **t-value** | **p-value** | **Evidence of Bias** |
| --- | --- | --- | --- | --- | --- | --- | --- |
| **Self-management behaviors** | | | | | | | |
| **Multiple TMFs** | 6 | 13.52 | 2.90 | 5.47 to 21.57 | 4.66 | 0.01 | Yes |
| **Self-management self-efficacy** | | | | | | | |
| **Multiple TMFs** | 14 | 0.52 | 1.41 | -2.56 to 3.59 | 0.37 | 0.72 | No |
| **Self-efficacy theory** | 8 | 0.80 | 1.73 | -3.42 to 5.03 | 0.47 | 0.66 | No |

**Note**: SE = Standard Error. A statistically significant intercept (*p* < 0.05) suggests the presence of funnel plot asymmetry and potential publication bias.
